# Heterogeneity in the Estimated Prevalence of Huntington’s Disease

**DOI:** 10.1101/2025.04.09.25325546

**Authors:** Alexander Thompson, Oliver Quarrell, Mark Strong

**Affiliations:** Yorkshire and Humber School of Public Health, Leeds, United Kingdom; School of Medicine and Population Health, University of Sheffield, Sheffield, United Kingdom; Sheffield Children’s Hospital, Sheffield, United Kingdom

## Abstract

There is significant variation in estimates of Huntington’s Disease (HD) prevalence in different settings. Despite this heterogeneity, there continues to be much interest in quantitatively synthesizing prevalence estimates to produce global or regional pooled prevalence values. This systematic review was undertaken to describe and assess the sources of heterogeneity in estimated prevalence values, and to consequently evaluate the validity of pooled prevalence values produced. Observational studies from which a prevalence estimate (point or period) or cumulative incidence of HD could be calculated between 1993-2024 were sought from Medline and Embase databases. Features of included studies are described and evaluated, with sources of heterogeneity discussed. A meta-regression was conducted including predictor variables: continent, median age of population, number of years since 1993, case ascertainment method, and Healthcare Access and Quality Index score. A total of 43 studies met the criteria for inclusion in the review. Significant clinical and methodological heterogeneity between the included studies is described, including differences in case definitions and ascertainment methods, and in the estimates of disease burden calculated. There were differences in the estimated point prevalence between regions and population groups within regions, while the estimated point prevalence was shown to be increasing over time since 1993. Wide prediction intervals in the overall pooled point prevalence (95% prediction interval: 0.32 – 37.55 cases per 100,000), and the European pooled point prevalence (95% prediction interval: 1.64 – 19.18 cases per 100,000), indicate the scale of heterogeneity between studies and settings. Such heterogeneity precludes valid extrapolation of findings to settings in which HD prevalence is unknown, and greatly limits the utility of pooled prevalence values.

## Introduction

Huntington’s disease (HD) is a genetic neurological disorder with autosomal dominant inheritance.^1^ It produces a wide spectrum of clinical symptoms and signs, including motor, cognitive, and behavioral features.^2^ The *HTT* gene responsible for HD was identified and described in 1993.^3^ In HD patients there is an expanded CAG trinucleotide repeat within the gene, encoding an abnormally long and toxic polyglutamine sequence.^4^ Average CAG repeat lengths in the general population have been described as between 16-20.^5^ Intermediate alleles, which may expand to pathogenic range on transmission from parents to offspring, have between 27-35 CAG repeats. 36-39 CAG repeats are associated with incomplete penetrance of HD, while 40 or more CAG repeats invariably leads to disease.

Historically, the diagnosis of HD was solely clinical, with patients identified on the basis of motor abnormalities (typically chorea), and cognitive and neuropsychiatric features, in conjunction with a family history of HD.^2^ Since the identification of the mutant *HTT* gene in 1993, the clinical diagnosis typically includes genetic confirmation of the expanded CAG repeat sequence. Prior to genetic testing it is likely that *de novo* cases of disease in patients without a family history, thought to represent around 5-8% of diagnoses, were missed.^6^

In recent years much interest has been given to describing the burden of HD in different populations.^7–11^ HD can have a significant health and social care impact on both affected individuals and families, and the wider health service.^12–13^ The accurate description and prediction of changes in the burden of the disease in a population is therefore of public health significance. In epidemiological terms the burden of a disease can be described using different measures. In the HD literature, these have typically included point prevalence (the proportion of the population with the disease at a single point in time), period prevalence (the total number of individuals with a diagnosis at any time during a period, divided by the total population over that period), and cumulative incidence (the total number of new cases during a period, divided by the total population over that period). These provide related but distinct measures of disease burden. In this manuscript we use the term ‘prevalence’ broadly to describe disease burden in a population. Where specific estimates are referenced, we specify the measure of disease burden used where possible.

Meta-analysis refers to the statistical combination of results from multiple studies to produce a pooled estimate.^14^ While historically meta-analyses have been used to pool results of multiple randomized controlled trials to produce an overall estimate of treatment effect, in recent years the methodology has been increasingly applied to studies measuring the prevalence of disease.^15^ The summary value reported is a pooled prevalence estimate – a weighted average – of all of the prevalence estimates synthesized. Indeed, in a recent systematic review and meta-analysis the ‘global prevalence’ of HD was reported as 4.88 per 100,000 (95% confidence interval (CI): 3.38-7.06).^10^ However, the apparently high heterogeneity in the prevalence of HD (described by large I^2^ values) between different populations, and at different times, raises questions about the validity and meaningfulness of such a quantitative synthesis.^16^ Such levels of heterogeneity in the context of studies measuring treatment effects would ordinarily preclude statistical synthesis.^14^

High values of I^2^ are ubiquitous in meta-analyses of prevalence.^15^ Observational studies of disease prevalence typically have large sample sizes (often the whole population), resulting in precise estimates of prevalence with low sampling error. Since the I^2^ statistic is a proportional value describing the proportion of heterogeneity due to variance in underlying true prevalence, it follows that as the sampling error tends towards zero the I^2^ statistic tends towards 100%. This is irrespective of whether true variation is large or small in absolute terms. Consequently, caution has been advised in excluding pooled estimates on the basis of large I^2^ values alone.^17^ Prediction intervals have been suggested as a more conservative method of incorporating uncertainty into a meta-analysis where true heterogeneity is expected.^18^ A recent meta-analysis in Latin America produced a regional prediction interval, although the authors advised caution in interpretation since the interval covered a large range.^11^

Clinical heterogeneity in the estimated prevalence of HD in different populations results from multiple factors. The underlying genetics of the population under investigation has been shown to be correlated with disease prevalence, including: mean CAG repeat length, presence of different genetic haplotypes, and CCG polymorphisms.^19–21^ Further, while HD can occur at almost any age the majority of patients develop HD between 35-55 years. Therefore, the age-distribution of the population has implications for the prevalence of disease. Since the estimated prevalence of HD is purportedly increasing in parts of the world, the date of any prevalence studies may impact on prevalence values reported.^22^

Methodological heterogeneity may also impact on estimates of disease prevalence. Countries with less developed healthcare services, or countries with higher stigma around disease diagnosis, may diagnose fewer cases.^23^ Additionally, study designs benefitting from an active case finding component, in which secondary cases are actively sought, may capture more cases than designs dependent on administrative data alone.^24^ Recent studies have made use of administrative healthcare records to identify HD cases.^22^ In the absence of case validation, this may lead to the inclusion of miscoded or misdiagnosed HD cases.^25^

Factors contributing to heterogeneity in the estimated prevalence can be elucidated by means of a meta-regression. A meta-regression regresses the outcome variable on various predictor variables associated with each study. Its use to explore heterogeneity is common in meta-analysis designs.^26^

In this study we aim to explore qualitatively and quantitatively, the clinical and methodological heterogeneity between included studies. Heterogeneity is visualized in a forest plot of included studies and summarized by I^2^ values and prediction intervals. Contributing factors to heterogeneity are described and quantitatively evaluated in a meta-regression analysis.

## Methods

The methodology followed the Preferred Reporting Items for Systematic Reviews and Meta-Analysis (PRISMA) protocols checklist. This systematic review is registered on Prospero: CRD42024605294.

### Study Selection

We conducted a systematic search of Medline and Embase databases using index terms specific to the prevalence and epidemiology of HD. The search was produced using minor alterations to a search conducted in a previous review (supplementary material).^10^ Two authors (AT/OQ) conducted all stages of the literature review independently. Studies were progressed through the title and abstract screening stage to full record review if they examined either case numbers or prevalence of HD in a defined geographical area. All discrepant selections at title and abstract screening were progressed to the next stage of review for both authors.

At full record review studies were included if: (1) there was sufficient information to calculate a measure of disease burden (point prevalence; period prevalence; cumulative incidence) and binomial confidence interval (at least 2/3 of: the proportional value case numbers/population size, population size, or case numbers), (2) the date of the measure of disease burden was 1993 or later, (3) the full paper was available in English, (4) the population size was at least 100,000, and (5) values reported were based on observational data in a defined geographical region or a representative sample of that region. Papers were excluded if they were a conference abstract or if they reported a mathematically modelled value. The 1993 date limit was set since this followed the identification of the mutant *HTT* gene, after which time testing for the gene became more routine in clinical practice. The 100,000-population size minimum was set since, given the rarity of HD, reports of disease burden in populations below this size would provide unreliable estimates. This approach is similar to previous reviews.^8^

Review papers and included articles had their references searched for further papers. Discrepancies in final study selection were discussed between three authors (AT/OQ/MS) until consensus on inclusion was reached. Referencing software Zotero 6.0.37 was used.

### Data Extraction

Manual data extraction for all studies was undertaken independently by two authors (AT/OQ). Data extracted included: date of study, number of cases, population size, case ascertainment method, case definition, and location (including continent). Where one of the ‘number of cases’ or the ‘total population size’ was not available, this was extrapolated from back-calculation. Where back-calculation was not possible, but there were reliable external sources of population size estimates (e.g. census records), these external sources were used. For studies presenting multiple point prevalence values at different time points, the case numbers and population size at each time point was extracted. Studies were grouped into three categories for the estimate of HD burden: point prevalence, period prevalence, or cumulative incidence. Case ascertainment methods were broadly categorized into ‘passive’ and ‘passive and active case finding’.

Data extracted was supplemented with online searches for the median age of the population studied and the Healthcare Access Quality Index (HAQI) score of the country. The HAQI score is an index measure which scores countries on a range from 0 – 100 based on both the quality of the healthcare provided and the ease of access for the general population. Where the population of interest was a region within a country, the median age for that country was applied. Any discrepancies in data extraction were discussed between all authors until consensus reached. Study details are available in the supplementary material.

### Risk of Bias

We assessed the risk of bias of each study using the Joanna Brigg’s Institute (JBI) critical appraisal tool for prevalence studies.^27^ This tool scores studies on their sampling, diagnostic, and statistical methodologies. Scores for each study are available in the supplementary material.

### Data Analysis

All studies were included within the comparative qualitative and descriptive analysis. For the quantitative data synthesis (meta-analysis and meta-regression), only a smaller subsection of these studies were included. Studies were included in the quantitative data synthesis if: (1) the estimate of disease burden calculated was a point prevalence, and (2) it was the most recent study within that region. Only including the most recent studies avoided the issue of patients being ‘counted twice’.

Data analysis and synthesis was undertaken using the ‘meta’ package in R version 2023.12.1+402. Studies are visually presented in a forest plot grouped by region. Studies were synthesized using a random effect meta-analysis (generalized linear mixed model (GLMM)) and 95% prediction intervals were produced. Prediction intervals describe the range of values within which 95% of similar studies are expected to lie. They are more conservative than confidence intervals since they incorporate study heterogeneity into the range described. Where no heterogeneity is present, prediction intervals coincide with confidence intervals.^28^ Prediction intervals for regional subgroups are only presented where n ≥ 10.^14^

The meta-regression was conducted using predictor variables agreed between authors *a priori.* These included: continent of study, number of years since 1993, HAQI score, median age of population, and case ascertainment method. As discussed in the introduction, these variables have been shown to be related to overall HD prevalence within a population. Unlike conventional regression analysis, meta-regression incorporates two additional error terms to account for sampling error in studies, and between-study heterogeneity. Meta-regression was conducted using the weighted least squares method.

### Sensitivity Analysis

The meta-analysis and meta-regression were repeated with differences in the definition of geographical regions (using the world bank classification) and excluding studies relying on administrative diagnostic codes for HD diagnosis.

## Results

### Literature Search

Medline and Embase databases were searched on 28^th^ October 2024 (Fig. 1). A total of 3092 citations were identified, 2409 in Embase and 684 in Medline. 673 of these records were excluded as duplicates. Titles were examined for relevance to HD prevalence, with 2000 records excluded at this stage. The abstracts of 419 records were reviewed, with 288 excluded. 131 records had their full texts examined, with 93 records excluded. Main reasons for exclusion included: insufficient information to calculate a prevalence value and binomial confidence interval (n=44), paper not available in English (n=15), and the paper being an abstract only (n=13). Other reasons included: paper being a review (n=9), a pre-1993 prevalence date (n=5), estimate of disease burden based on previously published data (n=5), population size <100,000 (n=1), and calculated prevalence based on a non-representative sample (n=1). Review papers and included records had their references searched for papers not identified through the search strategy, yielding five additional citations. A total of 43 records were included for analysis. A paper using insurance records in the USA was excluded from our analysis since the author’s acknowledged it was not a representative sample of the population.^29^ A study on a cluster of HD cases in the Minas Gerais state in Brazil was excluded since the population size was <100,000.^30^ Vishnevetsky et al’s 2023 study, while focused on juvenile HD in Peru, also reported numbers of adult HD patients and so was included.^31^ Details of the included studies are shown in table 1.

**Figure 1:**
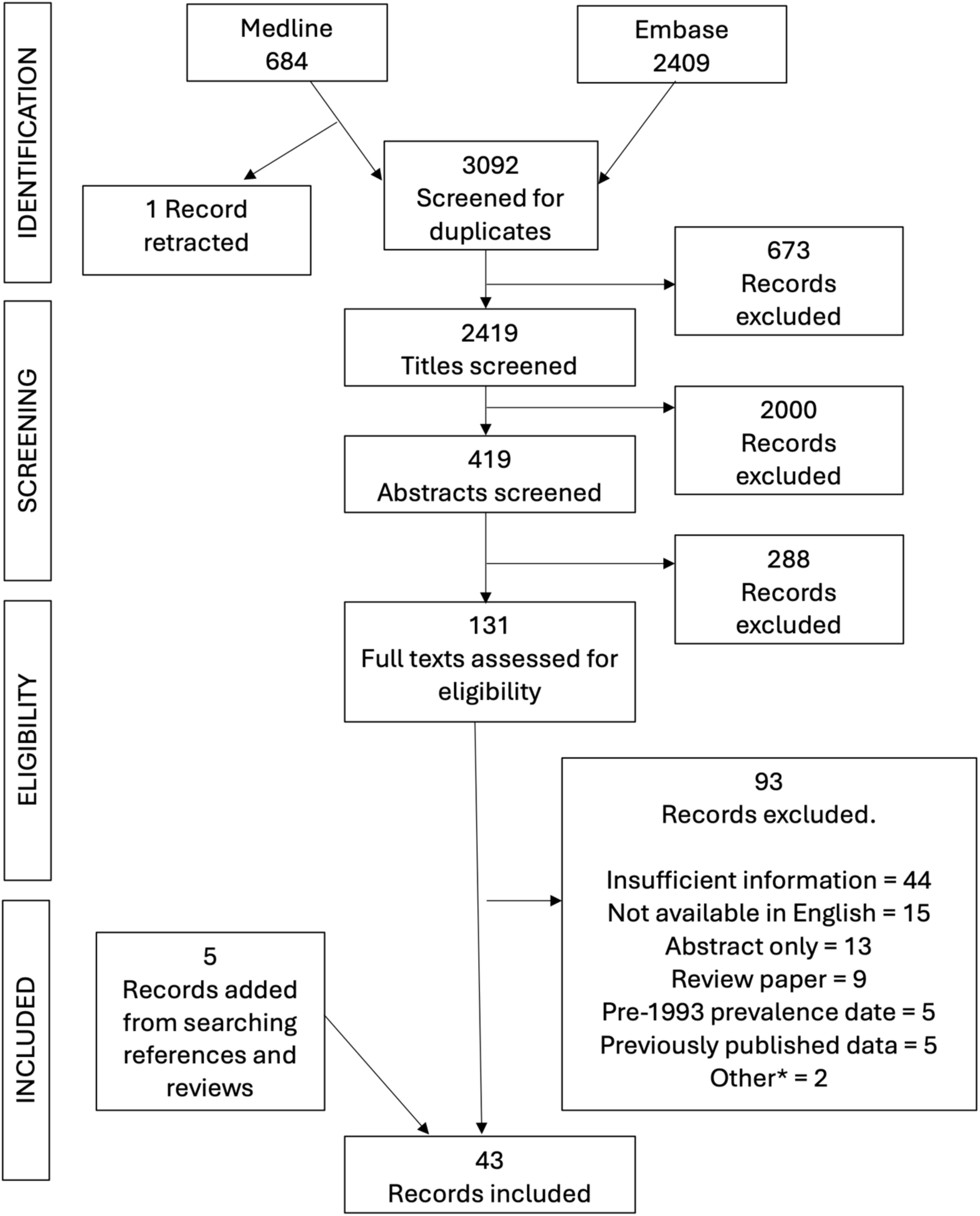
Flowchart of screening and selection process of final papers for inclusion in the systematic review. *Other= population size <100,000 (n=1) and a non-representative sample of a geographical region (n=1).

**Table 1:** Huntington’s Disease Prevalence Studies.

| **Table 1: Huntington's Disease Prevalence Studies** |  |  |  |  |  |  |  |  |
| --- | --- | --- | --- | --- | --- | --- | --- | --- |
| **Region** | **Prevalence date** | **Sources of cases** | **Diagnostic criteria** | **Population size** | **Number of cases** | **Cases per 100,000 population (95% CI)** | **Estimate Type** | **Reference** |
| **Europe** |  |  |  |  |  |  |  |  |
| Croatia | 2002 | Unspecified | Symptomatic + genetic confirmation | 4492049 | 44 | 0.98 (0.73, 1.32) | Unclear | Hećimović *et al.* (2002)<sup>32</sup> |
| Cyprus | 2015 | MR, ACF | Symptomatic + genetic confirmation | 840517 (calculated) | 39 | 4.64 (3.38, 6.36) | Point prevalence | Demetriou *et al.* (2017)<sup>33</sup> |
| Denmark | 2014 | DR, ACF | On disease register + clinically manifest | 5630000* | 329 | 5.84 (5.24, 6.51) | Point prevalence | Gilling *et al.* (2017)<sup>34</sup> |
| Finland | 2010 | MR | Administrative read code | 5377358 | 114 | 2.12 (1.76, 2.55) | Point prevalence | Sipilä *et al.* (2015)<sup>35</sup> |
| Finland | 2020 | MR | Administrative read code | 5500000 | 142 | 2.58 (2.19, 3.04) | Point prevalence | Sipilä and Majamaa (2023)<sup>36</sup> |
| Germany | 2015 – 2016 | IR | Administrative read code | 3325638 | 308 | 9.26 (8.28, 10.36) | Period prevalence | Ohlmeier *et al.* (2019)<sup>37</sup> |
| Greece | 2008 | Lab | Symptomatic + genetic confirmation | 10964020 | 594 | 5.42 (5, 5.87) | Cumulative incidence (1995 – 2008) | Panas *et al.* (2011)<sup>38</sup> |
| Iceland | 2007 | MR, ACF | Symptomatic + family history or genetic confirmation | 311114 | 3 | 0.96 (0.18, 2.98) | Point prevalence | Sveinsson *et al.* (2012)<sup>39</sup> |
| Italy (Ferrara) | 2014 | MR | Symptomatic + genetic confirmation | 354673 | 15 | 4.23 (2.5, 7.04) | Point prevalence | Carrassi *et al.* (2017)<sup>40</sup> |
| Italy (Molise) | 2013 | MR, DR, ACF | Clinical phenotype | 313341 | 34 | 10.85 (7.72, 15.21) | Point prevalence | Squitieri *et al.* (2015)<sup>41</sup> |
| Italy (South Sardinia and Cagliari) | 2017 | MR, Lab, ACF | Symptomatic + genetic confirmation | 785785 | 47 | 5.98 (4.48, 7.97) | Point prevalence | Muroni *et al.* (2020)<sup>42</sup> |
| Malta | 1994 | MR, ACF | Unspecified | 339173 | 40 | 11.79 (8.62, 16.1) | Point prevalence | Gassivaro *et al.* (1999)<sup>43</sup> |
| Northern Ireland | 2001 | DR, ACF | Symptomatic + ≥36 CAG repeats | 1698113 (calculated) | 180 | 10.6 (9.16, 12.27) | Point prevalence | Morrison *et al.* (2010)<sup>44</sup> |
| Russia (Vladimir Oblast) | 1994 – 1999 | MR, DR | Clinical + genetic confirmation | 1622900 | 31 | 1.91 (1.34, 2.72) | Period prevalence | Baryshnikova *et al.* (2002)<sup>45</sup> |
| Scotland (North) | 2020 | MR | Symptomatic + ≥36 CAG repeats | 893440 | 130 | 14.55 (12.25, 17.28) | Point prevalence | Kounidas *et al.* (2021)<sup>25</sup> |
| Slovenia | 2006 | MR, DR, ACF | Symptomatic + >36 CAG repeats | 2011614 | 104 | 5.17 (4.26, 6.27) | Point prevalence | Peterlin *et al.* (2009)<sup>46</sup> |
| Spain (Navarre) | 2017 | MR, ACF | Clinical + >35 CAG repeats or positive<br>family history | 647554 | 32 | 4.94 (3.48, 7) | Point prevalence | Vicente *et al.* (2021)<sup>47</sup> |
| Sweden | 2018 | MR | Administrative<br>read code | 10230185 | 1039 | 10.16<br>(9.56, 10.79) | Point<br>prevalence | Furby *et al.*<br>(2023)<sup>48</sup> |
| Sweden<br>(Jamtland) | 2015 | MR | Administrative<br>read code | 126765 | 28 | 22.1<br>(15.15, 32.06) | Point<br>prevalence | Roos *et al.*<br>(2017)<sup>49</sup> |
| Sweden<br>(Uppsala) | 2015 | MR | Administrative<br>read code | 348942 | 17 | 4.9<br>(2.98, 7.87) | Point<br>prevalence | Roos *et al.*<br>(2017)<sup>49</sup> |
| UK | 2018 | MR | Administrative<br>read code | 2593243 | 239 | 9.22<br>(8.12, 10.46) | Point<br>prevalence | Furby *et al.*<br>(2022)<sup>22</sup> |
| UK | 2010 | MR | Administrative<br>read code | 4683669 | 435 | 9.29<br>(8.45, 10.2) | Point<br>prevalence | Douglas *et al.* (2013)<sup>50</sup><br>and Evans *et al.* (2013)<sup>51</sup> |
| UK | 2011 | MR | Administrative<br>read code | 2964386 | 177 | 5.97<br>(5.15, 6.92) | Point<br>prevalence | Sackley *et al.*<br>(2011)<sup>52</sup> |
| Wales (South<br>East) | 1994 | DR, ACF | Clinical +<br>genetic | 1393900 | 86 | 6.17<br>(4.99, 7.63) | Point<br>prevalence | James *et al.*<br>(1994)<sup>53</sup> |
| **North**<br>**America** |  |  |  |  |  |  |  |  |
| Canada<br>(Alberta) | 2018 –<br>2019 | MR | Administrative<br>read code | 3183874 | 297 | 9.33<br>(8.32, 10.45) | Point<br>prevalence | Shaw *et al.*<br>(2022)<sup>12</sup> |
| Canada<br>(British<br>Columbia) | 2012 | MR, ACF | Symptomatic<br>+ Genetic<br>Confirmation<br>or Strictly<br>Clinical | 4609659 | 631 | 13.69<br>(12.66, 14.8) | Point<br>prevalence | Fisher *et al.*<br>(2014)<sup>54</sup> |
| US<br>(Navajo<br>Nation) | 2006 | MR | Administrative<br>read code | 217158 | 0 | 0<br>(0, 2.14) | Point<br>prevalence | Gordon *et al.*<br>(2016)<sup>55</sup> |
| **South America** |  |  |  |  |  |  |  |  |
| Brazil | 2016 | Lab, ACF | Symptomatic + molecular diagnosis | 11297297 (calculated) | 209 | 1.85 (1.62, 2.12) | Point prevalence | Castilhos *et al.* (2019)<sup>56</sup> |
| Chile | 2019 | MR, ACF | Symptomatic + >35 CAG repeats or positive family history | 19107216 | 138 | 0.72 (0.61, 0.85) | Point prevalence | Solís-Añez *et al.* (2023)<sup>57</sup> |
| Mexico (Mexico City) | 2008 | MR | Clinical or molecular diagnosis | 7950000 (calculated) | 318 | 4 (3.58, 4.47) | Cumulative incidence (1975 – 2008) | Alonso *et al.* (2009)<sup>58</sup> |
| Peru | 2000 – 2018 | MR | Genetically confirmed | 32203944* | 475 | 1.47 (1.35, 1.61) | Period prevalence | Vishnevetsky *et al.* (2022)<sup>31</sup> |
| Venezuela | 2006 | Lab | Symptomatic + expanded *HTT* allele | 27800000 (calculated) | 88 | 0.32 (0.26, 0.39) | Cumulative incidence (1988 – 2006) | Paradisi *et al.* (2008)<sup>59</sup> |
| **Asia** |  |  |  |  |  |  |  |  |
| Israel | 2018 | IR | Administrative read code | 1580816 | 69 | 4.36 (3.44, 5.53) | Point prevalence | Gavrielov-Yusim *et al.* (2021)<sup>60</sup> |
| Oman | 2019 | MR, ACF | Clinical or genetic confirmation | 556731 | 41 | 7.36 (5.4, 10.02) | Point prevalence | Squitieri *et al.* (2020)<sup>61</sup> |
| Japan (San-In area) | 1993 | MR | Symptomatic + expanded *HTT* allele | 1387000 | 9 | 0.65 (0.32, 1.25) | Point prevalence | Nakashima *et al.* (1996)<sup>62</sup> |
| South Korea | 2013 | IR | Administrative read code | 51141463 | 197 | 0.39<br>(0.33, 0.44) | Point prevalence | Kim *et al.* (2015)<sup>63</sup> |
| South Korea | 2019 | IR | Administrative read code | 51849861 | 1521 | 2.93<br>(2.79, 3.08) | Cumulative incidence (2010 – 2019) | Lee *et al.* (2023)<sup>64</sup> |
| Taiwan | 2007 | IR | Administrative read code | 23000000 | 97 | 0.42<br>(0.35, 0.51) | Point prevalence | Chen *et al.* (2010)<sup>65</sup> |
| **Oceania** |  |  |  |  |  |  |  |  |
| Australia (New South Wales) | 1996 | DR, MR, ACF | Symptomatic + positive DNA test or positive family history | 6038696 | 380 | 6.29<br>(5.69, 6.96) | Point prevalence | McCusker *et al.* (2000)<sup>66</sup> |
| Australia (Victoria) | 1999 | DR, Lab | Unspecified | 4736000 | 382 | 8.07<br>(7.3, 8.92) | Point prevalence | Tassicker *et al.* (2009)<sup>67</sup> |
| **Africa** |  |  |  |  |  |  |  |  |
| Cameroon | 2012 – 2014 | MR | Unspecified | 3380276 | 2 | 0.06<br>(0, 0.23) | Period prevalence | Cubo *et al.* (2017)<sup>68</sup> |
| Morocco (Rabat-Salé) | 2014 | MR | Symptomatic + genetic confirmation | 2000000 | 21 | 1.05<br>(0.68, 1.62) | Unclear | Bouhouche *et al.* (2015)<sup>69</sup> |
| South Africa | 2014 | Lab | Referred for testing + HTT expansion | 51207283<br>(calculated) | 384 | 0.75<br>(0.68, 0.83) | Cumulative incidence (1995 – 2014) | Baine *et al.* (2016)<sup>70</sup> |

**DR: Disease Register, MR: Medical Records, ACF: Active Case Finding, IR: Insurance Records. \*Taken from World Bank population estimates**

### Study Heterogeneity

Most included studies were undertaken in Europe (24/43; ∼56%). Asia and South America had six and five included studies respectively, Africa and North America both had three included studies, while Oceania had only two included studies.

There was variation in how cases were sourced. Most studies used medical records for at least part of their case sourcing (27/43; ∼63%). A HD register was used in seven included studies. Five studies used diagnostic codes from insurance records, while six studies included records from genetic laboratories. 14 studies benefitted from an active case finding component, typically searching for secondary cases within families.^61^ In one study the source of cases was unclear.^32^

There was variation in how HD cases were defined. Just over half of included studies incorporated clinical features with genetic confirmation into their case definition (23/43; ∼53%). Thirteen studies used administrative codes alone to identify HD diagnoses – of these only two detailed attempts to validate the diagnosis. Case validation in these studies involved clinical review of medical records with HD diagnostic read codes to confirm the diagnosis.^35,49^ An insurance-based study in Israel did limit their source of administrative records to either a neurologist diagnosis, or a chronic diagnosis, to improve the specificity of their case identification.^60^ In three studies the case definition was unclear.

Most studies included used the whole population size to describe the ‘population at risk’. However, an administrative based study in the UK, an insurance-based study in Israel, and a study in Croatia, included only adults aged ≥18 years of age,^22,32,60^ while a Canadian study included only adults ≥21 years.^12^ The prevalence of juvenile HD is known to be far lower than adult-onset.^50^

There was differential inclusion of incident and/or prevalent HD cases in the numerator of the disease burden estimates between studies, as well as whether efforts were made to exclude deceased patients on the prevalence date. A point prevalence value could be calculated in most studies (34/43; ∼79%). Period prevalence could be calculated in four of the studies. The period prevalence range varied between two and 18 years. Most of the remaining studies (5/43; ∼12%) appeared to include only incident cases diagnosed over the study period. The range of study periods for which cumulative incidence rates were reported was between 9-33 years. In two studies it was unclear how the prevalence value reported had been produced.^32,69^

There was variation in study quality as assessed using the JBI critical appraisal tool.^27^ Relatively few studies dropped points for their sample frame (6/43), sampling method (8/43) or sample size (42/43). Larger numbers of studies dropped points on the validity of methods used to diagnose the condition (29/43) – for instance, relying on administrative codes alone – and whether they clearly stated the numerator and denominator in their reporting of results (33/43).

### Meta-analysis

Study heterogeneity can be visualized in the forest plot in figure 2. Only studies for which a point prevalence value could be calculated were included in the quantitative synthesis and forest plot. Additionally, older studies were excluded where a more recent study in a geographical area was available. In the UK, Furby et al.’s 2022 study was preferred to Kounidas et al.’s 2021 study in Scotland, despite it having an older prevalence date, since it covered a broader and more representative sample of the whole UK population.^22,46^ Studies in other devolved nations in the UK were excluded since the CPRD database used in Furby et al.’s study includes practices based in all devolved nations. The final number of populations included in the quantitative synthesis was n=24.

Significant heterogeneity was present both within and between regional subgroups. The overall pooled 95% prediction interval for all studies ranged between 0.32 to 37.55 HD cases per 100,000. Large I^2^ values were reported for all regional subgroups, with a wide 95% prediction interval in Europe (1.64, 19.18). Prediction intervals in other regions are not presented since the subgroup sample size was insufficient. Given the significant heterogeneity present we do not report an overall pooled point prevalence value, nor pooled point prevalence values for the regional subgroups. The 95% confidence interval of the pooled prevalence value (2.19, 5.46) is presented as the checked lines on the forest plot. Only 10/24 included studies had 95% CIs that overlapped with the 95% CI of the pooled estimate, emphasizing the heterogeneity between study findings.

There is a large range of point prevalence values reported within European populations. Studies in Iceland and Finland reported low point prevalence values (0.96 and 0.98 per 100,000 respectively), ^32,36^ while higher point prevalence values were reported in small studies in Italy and Malta, ^41,43^ and a large administrative-based study in Sweden.^48^

The other continents had relatively few studies for inclusion. Asian studies included three in South-East Asia (Japan, South Korea, Taiwan) with similar reported point prevalence values (0.39-0.65 per 100,000), ^62–64^ while two Asian studies in the Middle Eastern region had higher point prevalence values reported (4.36-7.36 per 100,000).^60–61^ In North America a study based on the Navajo Nation found no cases of HD, ^55^ while studies in Canada found high point prevalence values similar to those reported in some of the higher prevalence areas in Europe.^12,54^

**Figure 2:**
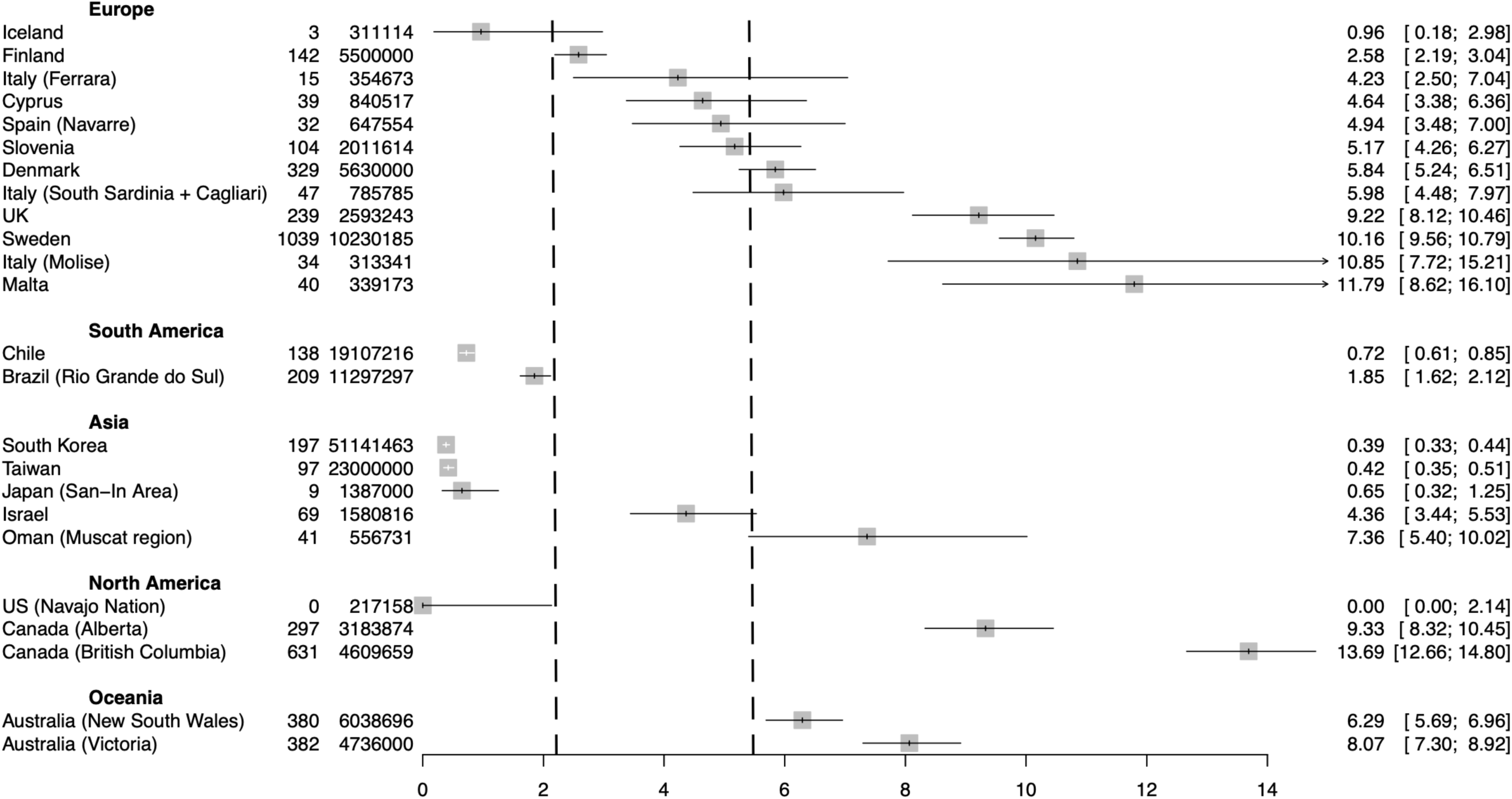
Forest plot presenting HD point prevalence estimates grouped by region. Error bars are 95% confidence intervals. Checked lines are the lower and upper 95% confidence limits for the pooled point prevalence estimate.

**Figure 3:**
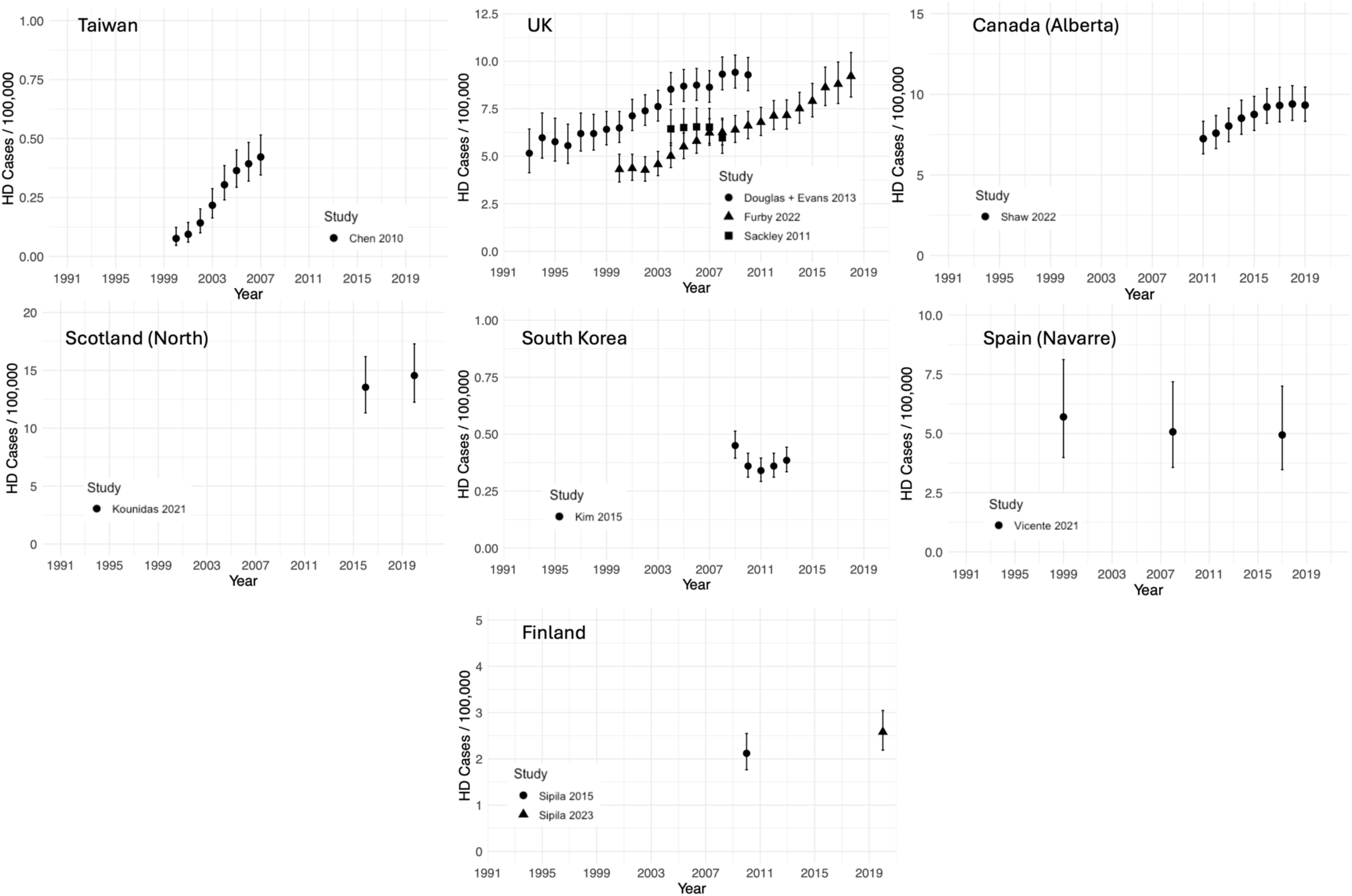
Point prevalence estimates over time. Error bars are 95% confidence intervals.

### Meta-regression

Table 2 presents the meta-regression analysis. Both Asian and South American based studies had a significantly reduced point prevalence compared to the European reference group. Prevalence in the Asian based studies were estimated at about a quarter of the European reference group, while South American studies were around a twentieth. Studies in Oceania and North America were not statistically significantly different compared to Europe.

**Table 2:** Meta-regression Results.

| **Table 2: Meta-regression Results** |  |  |  |
| --- | --- | --- | --- |
| **Variables** | **Exponentiated Coefficients (CI, 95%)** | **Standard Error** | **P - value** |
| Continent |  |  |  |
| Europe | 1 (ref) | NA | NA |
| Asia | 0.26 (0.09, 0.78) | 0.51 | 0.019 |
| South America | 0.05 (0.01, 0.3) | 0.86 | 0.003 |
| North America | 1.1 (0.37, 3.25) | 0.51 | 0.855 |
| Oceania | 4.45 (0.92, 21.5) | 0.74 | 0.062 |
| Median Age | 1 (0.91, 1.11) | 0.05 | 0.987 |
| Year post-1993 | 1.07 (1.01, 1.13) | 0.26 | 0.022 |
| Case Ascertainment Method |  |  |  |
| Passive | 1 (ref) | NA | NA |
| Passive and Active Case Finding | 1.97 (0.92, 4.24) | 0.36 | 0.078 |
| Healthcare Access and Quality Index (HAQI) score | 0.94 (0.85, 1.04) | 0.05 | 0.228 |
| Constant | 0.001 | 3.4 | 0.072 |
| $R^2$ | 0.62 | | |

Each year post-1993 was associated with approximately a 7% increase in the point prevalence of HD. The HAQI scores of the study countries was not associated with differences in the point prevalence of HD, and neither was the median age of the underlying population. While our model co-variates explained a significant proportion of the heterogeneity between studies (R^2^ = 0.62), nearly 40% of the heterogeneity between studies remained unexplained.

### Prevalence over time

For some geographical areas there were studies estimating point prevalence values at different points in time. Changes in prevalence over time in these areas are presented in figure 3.

The estimated point prevalence appears to have risen over time in: Taiwan, the UK, and the Alberta region of Canada.^12,22,50–52,65^ Four studies (the Douglas et al. and Evans et al. studies were combined to include both juvenile and adult-onset HD) using administrative primary care datasets are included within the UK region, with each study claiming to be based on a representative sample of the UK population. It is notable that in 2004 there is little to no overlap in the UK prevalence estimates between studies, despite apparent similarity of methods and large sample sizes. Further, while Furby et al.’s 2022 study is consistently lower than the other UK HD prevalence estimates presented, since this study actually reports a prevalence in only the ≥18-years-old adult population, we would have expected this study to estimate larger prevalence values than the other UK studies included. ^22^

There is little apparent change in estimated prevalence between the time points measured in: Scotland (Northern region), South Korea, Finland, and the Navarre region of Spain.^25,35,36,47,63^ To understand changes in the estimated prevalence of disease over time, it is important to consider changes in both the incidence and mortality of disease over the same period. Of the papers presented in figure 3 only Vicente et al.’s 2021 study presented contemporaneous incidence and mortality data.^47^ Five of the included studies did present data on incidence alone.^12,22,52,63,65^ Increasing point prevalence mirrors increasing incidence in Taiwan.^65^ However, in the UK and Canada apparent rises in the estimated point prevalence values occurred against a background of fluctuating incidence levels, indicating that improvements in patient longevity may have driven the changes.^12,22^

### Sensitivity Analysis

The meta-analysis and meta-regression were repeated with an alternative regional grouping (World Bank classification). We also repeated the meta-analysis and meta-regression excluding studies based on administrative data alone (supplementary material).

Significant heterogeneity remained, with large regional I^2^ values and wide overall and European (or ‘Europe and Central Asia’) 95% prediction intervals. Adjusting the region definitions to the World Bank classification meant that none of the adjusted meta-regression coefficients were statistically significant at the 5% level. When administrative code-based studies were excluded the adjusted meta-regression coefficients were grossly similar, with South American and Asian studies being associated with lower prevalence of HD, while each additional year post-1993 was associated with a 7% increase in HD prevalence. However, in this model increases in the HAQI score were associated with decreases in the prevalence of HD.

## Discussion

### Heterogeneity between regions

The large heterogeneity between studies in different regions is demonstrated by the wide overall pooled prediction interval incorporating all studies. The overall 95% prediction interval ranged between 0.32-37.55 HD cases per 100,000 – an over 100-fold difference in possible point prevalence values. While prediction intervals had been suggested as a mechanism for more conservatively synthesizing studies where true heterogeneity is expected,^17^ the overall prediction interval produced in the context of such heterogeneity is too wide to be meaningful. The adjusted meta-regression analysis demonstrated reduced HD point prevalence within both the South American and Asian continents, compared with the European reference group (around a quarter in Asian based studies; a twentieth in South American based studies). Lesser HD prevalence in Asian populations is widely recognized and likely reflects a combination of genetic factors: protective genetic haplotypes, reduced mean CAG repeat length in the general population, and CCG polymorphisms.^71^ The small sample of South American studies (n=2) limits conclusions based on this result.

### Heterogeneity within regions

Heterogeneity between studies in the same region is demonstrated by large regional I^2^ values (all >90%). The prediction interval for the European subgroup ranged between 1.64-19.48 HD cases per 100,000. This significant within region study heterogeneity indicates that ‘lines on a map’ may be a poor proxy for genetic similarity between populations within defined geographic regions. Indeed, Gordon et al.’s 2016 paper in the Navajo Nation in the US found no cases of HD, while papers based in Canada reported point prevalence values closer to the high prevalence populations in Europe.^12,54–55^ While these populations co-exist in the geographically defined North American continent, genetic differences between the different races studied will contribute to the differences in point prevalence estimates reported. Further, where HD burden is broken down by race, as in Baine et al.’s 2016 paper reporting HD cases in South Africa, racial differences are a much more significant predictor of genetic similarity, and consequently HD burden, than geographical area.^70^ An approach which pooled racially disparate populations into overall regional populations would lose important nuance in differences in HD prevalence between different racial groups. This is perhaps clearest in the ‘Asian’ studies within this systematic review. While studies based in South-East Asia reported largely similar point prevalence values (Taiwan, South Korea, and Japan),^62–63,65^ when combined with the Israel and Oman studies in the Middle East under the ‘Asia’ umbrella, the heterogeneity dramatically increases.^60–61^

### Heterogeneity over time

There is heterogeneity in point prevalence values observed at different times within our inclusion window. The adjusted meta-regression analysis demonstrated a positive association between the number of years since 1993 (the start of our inclusion window) and the prevalence date. On average, for each year after 1993 there was a 7% increase in the estimated HD point prevalence. Increasing estimates over time is also demonstrated in the included studies reporting point prevalence values at different time points (figure 3). In Taiwan, the UK, and the Alberta region of Canada, there are clear increases in the estimated point prevalence over time. Increasing estimates of point prevalence over time was not universal: studies in Spain and South Korea, did not demonstrate increases.^45,63^ Suggested factors driving increases in the estimated point prevalence include an increase in expected survival of HD patients from diagnosis, and reduced stigma around disease diagnosis.^23^ Additionally, where studies are repeated in the same region at different time points an increase in point prevalence may be noted due to improved case ascertainment over time (for instance, diagnosing secondary cases in families with established heterozygosity for the expanded *HTT* allele). Moreover, widening access to genetic testing may have facilitated further diagnoses.

While increases in disease prevalence can follow increases in disease incidence, Furby et al.’s 2022 paper in the UK reported unchanged HD incidence rates over the 19-year study period.^22^ It is also recognized that some under ascertainment on the prevalence date may occur due to the presence of symptomatic individuals who have not yet been diagnosed. Attempts to address this cause of under ascertainment have been made by using a prevalence date just before, or early in, the survey period.^44,66^ Patients symptomatic on the prevalence date but not diagnosed until later in the study period would therefore be included. We recommend that studies reporting a single estimated point prevalence value use remote prevalence dates where possible or if using estimates over time, include contemporaneous data on incidence and mortality.

### Heterogeneity in estimates of disease burden

There are significant differences in the estimates of HD burden that can be calculated from studies reporting HD cases. Of the 43 studies included within this systematic review, four could be used to produce estimates of period prevalence, while estimates of the cumulative incidence of HD cases could be calculated in five studies. Point prevalence values could be calculated in 34 studies. For valid quantitative synthesis, these distinctions in estimate types matter. Period prevalence values include all cases (prevalent and incident) during a time period, without excluding any cases that died during the study period. Consequently, the estimated number of cases is inflated (since deaths are not removed) and the prevalence value is also inflated. This is exemplified in Shaw’s 2022 Canadian study reporting yearly point prevalence values for the period 2014-2019, and a period prevalence value across the same six years.^12^ While the yearly point prevalence values range around 8 – 9 / 100,000, the period prevalence value reported over the same period is about a third higher at 12.15 / 100,000. Conversely, cumulative incidence values may provide either inflated or deflated approximations of point prevalence. Although case numbers are reduced since prevalent cases are not included, failing to exclude deceased patients will inflate case numbers reported. Consequently, whether or not a cumulative incidence estimate is greater or lesser than a point prevalence estimate depends on the length of the study period. Brief study periods, such as Bouhouche et al.’s 2022 Morocco based study will likely produce cumulative incidence values less than the ‘true’ point prevalence, since most cases in the region will be prevalent and therefore missed, while only few incident cases would be expected to have died during the study period.^69^ It is concerning that previous meta-analyses of HD prevalence have quantitatively synthesized studies reporting estimates of HD burden across all three categories described, without distinction or comment.^10,11^ We recommend that studies intending to report a measure of burden of HD make clear the estimate type reported.

### Heterogeneity in case ascertainment

There are significant differences between studies in how HD cases were ascertained. Thirteen of the included studies used administrative read codes alone to find HD cases. There is concern in the literature around the validity of using administrative codes for this purpose, which is dependent on the quality of the clinical coding.^47^ Indeed, inflation of case numbers due to false positives in administrative codes may be significant. Sipilä et al.’s 2015 study in Finland incorporated clinical review of medical records into their case ascertainment process.^35^ Of 399 records identified as having an HD diagnostic code, only 207 cases were validated as HD following chart review. Further, four UK based studies using administrative records from primary care databases all reported differing point prevalence estimates for the same time periods.^22,50–52^ While the rarity of HD makes traditional door-to-door survey methods for case finding impractical in most settings, it is important that the use of routine administrative data in case finding is rigorous and validated. Further attempts to assess and improve the validity of administrative data in epidemiological studies of HD prevalence should be a priority for future research. Incorporating multiple methods of ascertainment into the study design provides a mechanism for both validating cases (through triangulation of evidence), and also for identifying cases that would otherwise have been missed.^47,54^

### Heterogeneity in defining the population at risk

Differences in how the population at risk of HD is defined, may also lead to inflation of the estimated HD point prevalence in some contexts. Within this systematic review three papers included only cases of adult-onset HD, and therefore included only the adult population within the population at risk.^12,22,60^ Since juvenile onset HD makes up only a small fraction of overall HD cases, ‘adult-only’ point prevalence values would be expected to be higher than the estimated prevalence in the whole population.^72^

### Strengths and Limitations

This systematic review benefits from the large number of studies that have been undertaken since the identification of the mutant *HTT* allele in 1993. This enables meaningful evaluation of the different sources of heterogeneity between studies and settings. In comparison to previous reviews which speculate qualitatively on the contribution of different factors to heterogeneity,^7^ our meta-regression approach enables quantification of the relative contribution of different factors to HD prevalence.

However, caution must be advised in interpreting our meta-regression co-efficients. The reduced sample size for our quantitative analysis (n=24) means our regression model may be underpowered to find a true association where one exists, while model overfitting may also occur. Nevertheless, statistically significant predictors of the point prevalence of HD in our adjusted meta-regression model included a lesser point prevalence of HD in South American and Asian populations compared with the European reference population, and an increase in the point prevalence of HD in the years after 1993. These findings are epidemiologically plausible and are consistent with findings in other studies.^9–11,71^ Larger sample sizes may have been possible if we had been able to include non-English studies within this review. The lack of non-English studies also meant studies based in Europe made up a large proportion of our sample (24/43). This European bias further limits the use of overall pooled prevalence estimates, which would be more reflective of the European population group rather than the global population. Previous systematic reviews of HD prevalence that included studies published in other languages had larger numbers of studies for analysis.^8,11^

## Conclusion

This systematic review describes the significant heterogeneity between studies used to estimate the burden of HD in different settings. Sources of clinical heterogeneity include differences between regions and between different population groups within regions, and differences in HD prevalence over time. Sources of methodological heterogeneity include differences in case definitions and case ascertainment methodologies, as well as differences in the estimates of disease burden produced. Meta-analysis, in the context of such heterogeneity, produces invalid pooled prevalence values that lack meaning and are inappropriate for extrapolation to contexts in which HD prevalence is unknown. While prediction intervals have been suggested as a mechanism for incorporating heterogeneity in estimates of HD prevalence to produce a range of possible prevalence values in similar studies, the prediction intervals produced here are too wide to be meaningful for researchers and implementers of health policy.

## Data Availability Statement

The data supporting the findings of this study can be found in the supplementary material of this article.

## Acknowledgement

We are grateful to Sheffield Children’s Hospital Library for their support of this research.

## Author Roles

A. Conception of research; B. Literature Search; C. Data Extraction D. Statistical Analysis; E. Writing of first draft; F. Review and feedback.

AT: B, C, D, E

OQ: A, B, C, F

MS: A, D, F

## References

1. McColgan P, Tabrizi SJ. Huntington’s disease: a clinical review. Eur J Neurol 2018;25: 24–34

2. Bates G, Dorsey R, Gusella J, et al. Huntington disease. Nat Revi Dis Prim 2015;1: 15005

3. Chial, H. Huntington’s Disease: The Discovery of the Huntingtin Gene. Nature Education 2008;1:71

4. MacDonald M, Ambrose C, Duyao M, et al. A novel gene containing a trinucleotide repeat that is expanded and unstable on Huntington’s disease chromosomes. Cell 1993;72:971–983

5. Squitieri F, Andrew S, Goldberg Y, et al. DNA haplotype analysis of Huntington disease reveals clues to the origins and mechanisms of CAG expansion and reasons for geographic variations of prevalence. Hum Mol Genet 1994;3:2103–14

6. Almqvist E, Elterman D, MacLeod P, Hayden M. High incidence rate and absent family histories in one quarter of patients newly diagnosed with Huntington disease in British Columbia. Clin Genet 2001;60:198–205

7. Baig S, Strong M, Quarrell O. The global prevalence of Huntington’s disease: a systematic review and discussion. Neurodegener Dis Manag 2016;6:331–43

8. Rawlins M, Wexler N, Wexler A, Tabrizi S, Douglas I, Evans S, Smeeth L. The Prevalence of Huntington’s Disease. Neuroepidemiology 2016;46:144–53

9. Pringsheim T, Wiltshire K, Day L, Dykeman J, Steeves T, Jette N. The Incidence and Prevalence of Huntington’s Disease: A Systematic Review and Meta-analysis. Mov Disord 2012;27:1083–91

10. Medina A, Mahjoub Y, Shaver L, Pringsheim T. Prevalence and Incidence of Huntington’s Disease: An Updated Systematic Review and Meta-Analysis. Mov Disord 2022;37:2327–2335

11. Medina A, Pringsheim T, Gautreau S, Rivera-Duarte J, Amorelli G, Cornejo-Olivas M, Rossi M. Epidemiology of Huntington’s Disease in Latin America: A Systematic Review and Meta-Analysis. Mov Disord 2024

12. Shaw E, Mayer M, Ekwaru P, et al. Disease Burden of Huntington’s Disease (HD) on People Living with HD and Care Partners in Canada. J. Huntington’s Dis. 2022;11:179–193

13. Divino V, Dekoven M, Warner J, et al. The direct medical costs of Huntington’s disease by stage. A retrospective commercial and Medicaid claims data analysis. J. Med. Econ. 2013;16:1043–50

14. Deeks J, Higgins J, Altman D. Chapter 10: Analysing data and undertaking meta-analyses. Cochrane Handbook for Systematic Reviews of Interventions. Cochrane, 2023.

15. Migliavaca C, Stein C, Colpani V, et al. Meta-analysis of prevalence: I2 statistic and how to deal with heterogeneity. Res Synth Methods 2022;13:363–67

16. Strong M, Quarrell O. Prevalence and Incidence of Huntington’s Disease. Mov Disord 2023;38:1570–72

17. Pringsheim, T. Reply to Letter to the Editor: Prevalence and Incidence of Huntington’s Disease Comment on Medina, et al. (2022). Mov Disord, 2023;38:1572–73

18. Borenstein M, Higgins J, Hedges L, et al. Basics of meta-analysis: I2 is not an absolute measure of heterogeneity. Res Synth Methods 2017;8:5–18

19. Warby S, Montpetit A, Hadyen A, et al. CAG Expansion in the Huntington Disease Gene Is Associated with a Specific and Targetable Predisposing Haplogroup. Am J Hum Genet 2009;84:351–66

20. Chao TK, Hu J, Pringsheim T. Risk factors for the onset and progression of Huntington disease. Neurotoxicology 2017;61:79–99

21. Xu M, Wu ZY. Huntington Disease in Asia. Chin Med J 2015;128:1815–9

22. Furby H, Siadimas A, Rutten-Jacobs L, Rodrigues F, Wild E. Natural history and burden of Huntington’s disease in the UK: A population-based cohort study. Eur J Neurol 2022;29:2249–2257

23. Rawlins M. Huntington’s disease out of the closet? Lancet 2010;376:1372–3

24. Harper P. The epidemiology of Huntington’s disease. Hum Genet 1992;89:365–76

25. Kounidas G, Cruickshank H, Kastora S et al. The known burden of Huntington disease in the North of Scotland: prevalence of manifest and identified pre-symptomatic gene expansion carriers in the molecular era. J Neurol 2021:4170–77

26. Migliavaca C, Stein C, Colpani V, et al. How are systematic reviews of prevalence conducted? A methodological study. BMC Med Res Methodol 2020;20:96

27. Munn Z, Moola S, Riitano D, Lisy K. The development of a critical appraisal tool for use in systematic reviews addressing questions of prevalence. Int J Health Policy Manag 2014;13:123–28

28. IntHout J, Ioannidis J, Rovers M, Goeman J. Plea for routinely presenting prediction intervals in meta-analysis. BMJ Open 2016;6:e010247

29. Bruzelius E, Scarpa J, Zhao Y, Basu S, Faghmous J, Baum A. Huntington’s disease in the United States: Variation by demographic and socioeconomic factors. Mov Disord 2019;34:858–65

30. Agostinho L, da Silva I, Maia L, et al. A Study of a Geographical Cluster of Huntington’s Disease in a Brazilian Town of Zona da Mata, Minas Gerais State. Eur Neurol 2015;74:62–8

31. Vishnevetsky A, Cornejo-Olivas M, Sarapura-Castro E, et al. Juvenile-onset Huntington’s disease in Peru: A case series of 32 patients. Mov Disord Clin Pract. 2023; 10(2): 238–247.

32. Hećimović S, Klepac N, Vlasic J, et al. Genetic Background of Huntington Disease in Croatia: Molecular Analysis of CAG, CCG, and Δ2642 (E2642del) Polymorphisms. Hum Mutat 2002;20:233

33. Demetriou C, Heraclides A, Salafori C, Tanteles G, Christodoulou K, Christou Y, Zamba-Papanicolaou E. Epidemiology of Huntington disease in Cyprus: A 20-year retrospective study. Clin Genet 2018;93:656–664

34. Gilling M, Budtz-Jorgensen E, Boonen S, Lildballe D, Bojesen A, Hertz J, Sorensen S. The Danish HD Registry—a nationwide family registry of HD families in Denmark. Clin Genet 2017;92:338–341

35. Sipilä J, Hietala M, Siitonen A, et al. Epidemiology of Huntington’s disease in Finland. Parkinsonism Relat Disord 2015;21:46–9

36. Sipilä J, Majamaa K. Stable low prevalence of Huntington’s disease in Finland. Clin Park Relat Disord 2023;8:100198

37. Ohlmeier C, Saum KU, Galetzka W, Beier D, Gothe H. Epidemiology and health care utilization of patients suffering from Huntington’s disease in Germany: real world evidence based on German claims data. BMC Neurol 2019;19:318

38. Panas M, Karadima G, Vassos E, Kalfakis N, Kladi A, Christodoulou K, Vassilopoulos D. Huntington’s disease in Greece: the experience of 14 years. Clin Genet 2011;80:586–90

39. Sveinsson O, Halldórsson S, Olafsson E. An Unusually Low Prevalence of Huntington’s Disease in Iceland. Eur Neurol, 2012;68:48–51

40. Carrassi E, Pugliatti M, Govoni V, Sensi M, Casetta I, Granieri E. Epidemiological Study of Huntington’s Disease in the Province of Ferrara, Italy. Neuroepidemiology 2017;49:18–23

41. Squitieri F, Griguoli A, Capelli G, Porcellini A, D’Allesio B. Epidemiology of Huntington disease: first post-HTT gene analysis of prevalence in Italy. Clin Genet 2015:89;367–70

42. Muroni A, Murru M, Sechi M et al. Prevalence of Huntington’s disease in Southern Sardinia, Italy. Parkinsonism Relat Disord 2020;80:54–57

43. Gassivaro G, Buhagiar M, Cuschieri A, Viviani F. Huntington’s chorea (HD) in Malta: Epidemiology and origins. Int J Anthropol Ethnol 1999;14:115–125

44. Morrison P, Harding-Lester S, Bradley A. Uptake of Huntington disease predictive testing in a complete population. Clin Genet 2011;80:281–6

45. Baryshnikova N, Dadali E, Okuneva E et al. Hereditary Diseases of Nervous System in the Population of Vladimir Oblast. Genetika 2002;38:400–6

46. Peterlin B, Kobal J, Teran N, Flisar D, Lovrecic L. Epidemiology of Huntington’s disease in Slovenia. Acta Neurol Scand 2009;119:371–5

47. Vicente E, Ruiz de Sabando A, Garcia F, Gaston I, Ardanaz E, Ramos-Arroyo M. Validation of diagnostic codes and epidemiologic trends of Huntington disease: a population-based study in Navarre, Spain. Orphanet J Rare Dis 2021;16:77

48. Furby H, Moore S, Nordstroem AL, et al. Comorbidities and clinical outcomes in adult- and juvenile-onset Huntington’s disease: a study of linked Swedish National Registries (2002–2019). J Neurol 2023;270:864–876

49. Roos A, Wiklund L, Laurell K. Discrepancy in prevalence of Huntington’s disease in two Swedish regions. Acta Neurol Scand 2017;136:511–15

50. Douglas I, Evans S, Rawlins M, Smeeth L, Tabrizi S, Wexler N. Juvenile Huntington’s disease: a population-based study using the General Practice Research Database. BMJ Open 2013;3:e002085

51. Evans S, Douglas I, Rawlins M, et al. Prevalence of adult Huntington’s disease in the UK based on diagnoses recorded in general practice records. J Neurol Neurosurg Psychiatry 2013;84:1156–60

52. Sackley C, Hoppitt T, Calvert M, et al. Huntington’s Disease: Current Epidemiology and Pharmacological Management in UK Primary Care. Neuroepidemiology 2011;37:216–21

53. James C, Houlihan G, Snell R, et al. Late-onset Huntington’s disease: a clinical and molecular study. Age Ageing 1994;23:445–8

54. Fisher E, Hayden M. Multisource ascertainment of Huntington disease in Canada: Prevalence and population at risk. Mov Disord 2014;29:105–14

55. Gordon P, Mehal J, Rowland A, Cheek J, Bartholomew M. Huntington disease among the Navajo: A population-based study in the Navajo Nation. Neurology 2016;86:1552–3

56. Castilhos R, Santos J, Augustin M, et al. Minimal prevalence of Huntington’s disease in the South of Brazil and instability of the expanded CAG tract during intergenerational transmissions. Genet Mol Biol 2019;42:329–336

57. Solís-Ańez E, Salles P, Rojas N, Benavides O, Chana-Cuevas P. Huntington’s Disease in Chile: Epidemiological and Genetic Aspects. Neuroepidemiology 2023;57:176–184

58. Alonso ME, Ochoa A, Boll MC, et al. Clinical and genetic characteristics of Mexican Huntington’s disease patients. Mov Disord 2009; 24(13): 2012–2015.

59. Paradisi I, Hernández A, Arias S. Huntington disease mutation in Venezuela: age of onset, haplotype analyses and geographic aggregation. J Hum Genet 2008;53:127–135

60. Gavrielov-Yusim N, Barer Y, Martinec M, et al. Huntington’s Disease in Israel: A Population-Based Study Using 20 Years of Routinely-Collected Healthcare Data. J Huntingtons Dis 2021;10:469–77

61. Squitieri F, Maffi S, Al Harasi S, Al Salmi Q, D’Alessio B, Capelli G, Mazza T. Incidence and prevalence of Huntington disease (HD) in the Sultanate of Oman: the first Middle East post-HTT service-based study. J Neurol Neurosurg Psychiatry 2020;91:1359–60

62. Nakashima K, Watanabe Y, Kusumi M. Epidemiological and Genetic Studies of Huntington’s Disease in the San-in Area of Japan. Neuroepidemiology 1996;15:126–31

63. Kim HS, Lyoo CH, Lee PH, et al. Current Status of Huntington’s Disease in Korea: A Nationwide Survey and National Registry Analysis. Journal Mov Disorders 2015;8:14–20

64. Lee CY, Ro JS, Jung H, Kim M, Jeon B, Lee JY. Increased 10-Year Prevalence of Huntington’s Disease in South Korea: An Analysis of Medical Expenditure Through the National Healthcare System. J Clin Neurol 2023;19:147–55

65. Chen YY, Lai CH. Nationwide Population-Based Epidemiologic Study of Huntington’s Disease in Taiwan. Neuroepidemiology 2010;35:250–4

66. Mccusker E, Casse R, Graham S. Prevalence of Huntington disease in New South Wales in 1996. Med J Aust 2000;173:187–90

67. Tassicker R, Teltscher B, Trembath M, et al. Problems assessing uptake of Huntington disease predictive testing and a proposed solution. Eur J Hum Genet 2008;17:66–70

68. Cubo E, Doumbe J, Mapoure N, et al. The Burden of Movement Disorders in Cameroon: A Rural and Urban-Based Inpatient/Outpatient Study. Mov Disord Clin Pract 2017;4:568–573

69. Bouhouche A, Regragui W, Lamghari H, et al. Clinical and genetic data of Huntington disease in Moroccan patients. Afr Health Sci 2015;15:1232–8

70. Baine F, Krause A, Greenberg J. The Frequency of Huntington Disease and Huntington Disease-Like 2 in the South African Population. Neuroepidemiology 2016;46:198–202

71. Papanna B, Lazzari C, Rabottini M. The Prevalence of Huntington Disease in Asia Highlights Needs in Clinical, Genetic and Instrumental Diagnosis: A Systematic Review and Meta-Analysis. Psychiatr Danub 2022;34:13–23

72. Quarrell O, O’Donovan K, Bandmann O, et al. The Prevalence of Juvenile Huntington’s Disease: A Review of the Literature and Meta-Analysis. PLoS Curr 2012;4:e4f8606b742ef3

