## Supplementary Material for "Heterogeneity in the Estimated Prevalence of Huntington’s Disease"

**Embase Search**

1 exp Huntington chorea/di, ep, et [Diagnosis, Epidemiology, Etiology]
2 Huntington*.tw,kw.
3 exp incidence/ep [Epidemiology]
4 Incidence*.tw,kw.
5 exp prevalence/ep [Epidemiology]
6 prevalence*.tw,kw.
7 exp epidemiology/
8 epidemiology*.tw,kw.
9 3 or 4 or 5 or 6 or 7 or 8
10 1 or 2
11 9 and 10
12 Huntington chorea/ep
13 11 or 12
14 limit 13 to yr="1993 -Current"
15 limit 14 to animals
16 14 not 15

**Medline Search**

1 exp Huntington Disease/cl, di, ep, et [Classification, Diagnosis, Epidemiology, Etiology]
2 Huntington*.tw,kf.
3 1 or 2
4 exp Incidence/
5 Incidence*.tw,kf.
6 4 or 5
7 exp Prevalence/
8 prevalence*.tw,kf.
9 7 or 8
10 exp Epidemiology/cl, di, mt, sn [Classification, Diagnosis, Methods, Statistics & Numerical Data]
11 epidemiology*.tw,kf.
12 10 or 11
13 6 or 9 or 12
14 3 and 13
15 *Huntington Disease/ep [Epidemiology]
16 14 or 15
17 limit 16 to yr="1993 -Current"
18 limit 17 to animals
19 17 not 18

**Supplementary Table 1: Included Prevalence Studies**

| Study, year, location | Case Definition | Cases | Population | Data Source | Prevalence Date | Overall Calculated Prevalence (95% CI) |
|---|---|---|---|---|---|---|
| Alonso, 2009, Mexico (Mexico City) | (1) having a clinical (involuntary movements, emotional disturbance, and cognitive decline) or molecular diagnosis of HD and (2) being of Mexican mestizo origin | 318<br><br>Incident cases in Mexico City between 1973-2008. | Produced by back-calculation: N=(318/4)*100,000<br><br>7950000<br><br>All ages.<br><br>Unclear how/what they have used as the denominator to produce a 4 / 100,000 prevalence. | National Institute of Neurology and Neurosurgery. | 1973-2008<br><br>Cumulative incidence of Mexico City HD patients over a 36-year period. | 4 / 100,000 (3.58, 4.47) |
| Baine, 2016, South Africa | Referred and genetically tested. | 384<br><br>Incident cases in South Africa between 1995-2014. | Produced by back-calculation: N=51207283<br><br>All ages.<br><br>Denominator appears to be population size in 2011. | Reviewed database records for all patients referred for HD testing. | 1995-2014<br><br>Cumulative incidence of South Africa HD patients over a 20-year period. | 0.75/100,000 (0.68, 0.83) |
| Baryshnikova, 2002, Russia (Vladimir Oblast) | Clinical + genetic confirmation (although unclear if all patients received genetic confirmation) | 31<br><br>Appears to include incident and prevalent cases | 1,622,900<br><br>All ages.<br><br>Denominator appears to be population size in 1989. | Neurological hospitals, outpatient clinics, medical sanitary units, the medico-social examination | 1994-1999<br><br>Period prevalence in Vladimir Oblast | 1.91 / 100,000 (1.34, 2.72) |

| | | | | | | |
|---|---|---|---|---|---|---|
| | | between 1994-1999. | | bureau for the period from 1994 to 1999, and from the Medical Genetic Counseling register | between 1994-1999. | |
| Bouhouche, 2015, Morocco (Rabat-Salé) | Symptomatic + genetic confirmation | 21<br><br>Incident cases in Rabat-Salé between 2009-2014. | 2,000,000<br><br>All ages.<br><br>Unclear which year has been used for the population size estimate. | Outpatient Clinics | 2009-2014<br><br>Cumulative incidence of Rabat-Salé HD patients over a 6-year period. | 1.05 / 100,000 (0.68, 1.62) |
| Carrasi, 2017, Italy (Ferrara) | CAG expansion + unexplained extrapyramidal movement disorder | 15<br><br>Total cases alive on prevalence date. | 354,673<br><br>All ages.<br><br>Total population size on prevalence date. | Retrospective chart review from archives and outpatient records. Hospital discharge records and the Regional Record for Rare Diseases. | Dec. 31st 2014<br><br>Point prevalence. | 4.23 / 100,000 (2.5, 7.04) |
| Castilhos, 2019, Brazil (Rio Grande do Sul) | Symptomatic + molecular diagnosis. | 209<br><br>Total cases alive on prevalence date. | Produced by back-calculation:<br>n = (209/1.85)*100,000<br><br>11,297,297<br><br>All ages. | Diagnosed at the author's institution. | Dec 2016<br><br>Point prevalence. | 1.85 / 100,000 (1.62, 2.12) |

| | | | | | | |
|---|---|---|---|---|---|---|
| | | | Estimated population size in 2017 based on 2010 census. | | | |
| Chen, 2010, Taiwan | ICD-9 Code 333.4 | 97<br><br>Total cases alive on prevalence date. | 23,000,000<br><br>All ages.<br><br>Population size (of insurance database) on prevalence date. | Annual outpatient claims and hospitalization discharge claims from the National Health Insurance Database/ | 2007<br><br>Point prevalence. | 0.42 / 100,000 (0.35, 0.51) |
| Cubo, 2017, Cameroon | Clinical diagnosis based on chart review | 2<br><br>Prevalent and incident cases, unclear if adjusted for deaths. | 3,380,276<br><br>All ages.<br><br>Unclear which year has been used for the population size estimate. | Medical charts reviewed of patients on the registry of neurological diseases. | 2012-2014<br><br>Period prevalence in Doula Cameroon between 2012-2014. | 0.06 / 100,000 (0, 0.23) |
| Demetriou, 2017, Cyprus | Clinical and molecular diagnosis | 39<br><br>(58 patients minus 17 adult patients who died and 2 JHD patients who died)<br><br>Total cases alive on prevalence date. | Produced by back-calculation:<br>(n = (39/4.64)*100,000)<br><br>840,517<br><br>All ages.<br><br>End of 2015 population size. | Patients identified from the neurology and genetics referral centre. Additional case finding undertaken by contacting neurologists. | 31st Dec 2015<br><br>Point prevalence. | 4.64 / 100,000 (3.58, 6.01) |
| Douglas + Evans, 2013, UK | One or more diagnostic read codes for HD in medical record. | 435<br><br>Total cases alive on prevalence date. | 4683669<br><br>All ages. | The General Practice Research Database (GPRD) | 2010<br><br>Point prevalence. | 9.29 / 100,000 (8.45, 10.2) |

| | | | | | | |
|---|---|---|---|---|---|---|
| | | | Total population size of database. | | | |
| Fisher, 2013, Canada (British Columbia) | Strictly clinical or clinical + genetic confirmation | 631<br><br>Total cases alive on prevalence date (patients diagnosed >19yrs ago and with no update to record in last 5 years were assumed deceased). | 4,609,659<br><br>All ages.<br><br>Total estimated population size on prevalence date. | Medical and laboratory records. Surveys of clinicians and nursing homes. Detailed chart review. | April 2012<br><br>Point prevalence. | 13.69 / 100,000 (12.66, 14.8) |
| Furby, 2022, UK | Read code: F134.00 Huntington's chorea; Eu02200 Dementia in HD | 239<br><br>Total cases alive on prevalence date. | 2,593,243<br><br>>18s only.<br><br>Total population size of database. | GP records using CPRD GOLD. | 2018<br><br>Point prevalence. | 9.22 / 100,000 (8.12, 10.46) |
| Furby, 2023, Sweden | ICD-10 diagnostic code | 1039<br><br>Total cases alive on prevalence date. | 10,230,185<br><br>All ages.<br><br>Total 2018 Swedish population. | Swedish national patient registry. | Jan 1st 2018<br><br>Point prevalence. | 10.16 / 100,000 (9.56, 10.79) |
| Gassivaro, 1999, Malta | Clinical and genetic | 40<br><br>Total number of living cases. | 339,173<br><br>All ages.<br><br>Total population size Jan 31st 1993. | Neurogenetic clinic of St Luke's Hospital | 1994<br><br>Point prevalence. | 11.79 / 100,000 (8.62, 16.1) |

| | | | | | | |
|---|---|---|---|---|---|---|
| Gavrielov-Yusim, 2021, Israel | ICD-9 diagnostic code | 69<br><br>Total number of living cases. | 1,580,816<br><br>Adults only.<br><br>Total population size of adults in database. | Insurance records from MHS | 2018<br><br>Point prevalence. | 4.36 /100,000 (3.44, 5.53) |
| Gilling, 2017, Denmark | Presence on disease register. | 329<br><br>Manifest patients alive on prevalence date. | 5,630,000<br><br>All ages.<br><br>Denominator taken from data on total population of Denmark in 2014. | Danish Huntington's Disease Registry | June 2014<br><br>Point prevalence. | 5.84 / 100,000 (5.2, 6.45) |
| Gordon, 2016, US (Navajo Nation) | At least 2 inpatient or outpatient visits with the diagnosis. | 0<br><br>Total number of living cases. | 217,158<br><br>All ages.<br><br>Denominator taken from supplementary material – total population size on prevalence date. | The Indian Health Service National Patient Info reporting system. | July 1st 2006<br><br>Point prevalence. | 0 / 100,000 (0, 2.14) |
| Hecimovic, 2002, Croatia | Clinical diagnosis from neurologist and genetic confirmation | 44<br><br>Only included patients >18yrs of age who gave informed consent. Unclear if any juvenile cases were excluded.<br><br>Total number of living cases. | 4,492,049<br><br>Unclear on the date of the total population size used to calculate the prevalence value. | Unclear. | 2002<br><br>Point prevalence. | 0.98 / 100,000 (0.73, 1.32) |

| | | | | | | |
|---|---|---|---|---|---|---|
| James, 1994, Wales | Identified on locally maintained disease register | 86<br><br>Total number of living cases. | 1,393,900<br><br>All ages.<br><br>Total estimated population size in 1990. | Wales Huntington Disease register | 1st March 1994<br><br>Point prevalence. | 6.17 / 100,000 (4.99, 7.63) |
| Kim, 2015, Korea | Identified as having HD from health insurance database | 197<br><br>Total number of living cases. | 51,141,463<br><br>All ages.<br><br>Total number of patients on database. | National Health Insurance database | Dec. 2013<br><br>Point prevalence. | 0.39 / 100,000 (0.33, 0.44) |
| Kounidas, 2021, Scotland | Patient living in North of Scotland on 01/01/2020 with 36 or more CAG repeats and with manifest disease. | 130<br><br>Total number of living cases. | 893,440<br><br>All ages.<br><br>Total population size of area for 2019. | NHS Grampian genetic department laboratory and clinic records. | 01/01/2020<br><br>Point prevalence. | 14.55 / 100,000 (12.25, 17.28) |
| Lee, 2023, South Korea | Diagnostic code G10 of ICD-10. | 1521<br><br>Total number of incident cases in South Korea over a 10 year period. | 51327916<br><br>All ages.<br><br>Population size mid-way through the study period (2014). | National Health Insurance Database | 2019<br><br>Cumulative incidence of South Korea HD patients over a 10-year period. | 2.96 / 100,000 (0.33, 0.44) |
| McCusker, 2000, Australia (New South Wales) | Definite HD diagnosis + family history or positive genetic test. Or, signs of HD at | 380<br><br>Total number of living cases. | 6,038,696<br><br>All ages. | Huntington's Disease Register, hospital records, questionnaire sent to neurologists and | 06/08/1996<br><br>Point prevalence. | 6.29 / 100,000 (5.69, 6.96) |

| | | | | | | |
|---|---|---|---|---|---|---|
| | prevalence date and later received a definite diagnosis. | | Total population size on prevalence date. | geneticists in the region. | | |
| Morrison, 2010, Northern Ireland | Symptomatic and genetic confirmation. | 180<br><br>Total number of patients alive on prevalence date. | Produced by back-calculation:<br>(n= (180/10.6)*100,000)<br><br>1,698,113<br><br>All ages.<br><br>Total population size on prevalence date. | Huntington's Disease Register. | Jan 1st 2001<br><br>Point prevalence. | 10.6 / 100,000 (9.16, 12.27) |
| Muroni, 2020, Italy (Cagliari and South Sardinia) | Symptomatic and genetic confirmation. | 47<br><br>Total number of patients alive on prevalence date. | 785785<br><br>All ages.<br><br>Total population size on prevalence date. | Contacted laboratories and medical clinics in the region, and contacted local physicians. | Dec 31st 2017<br><br>Point prevalence. | 5.98 / 100,000 (4.48, 7.97) |
| Nakashima, 1993, Japan (San-In Area) | Symptomatic and genetic confirmation. | 9<br><br>Total number of patients alive on prevalence date. | 1387000<br><br>All ages.<br><br>Total population size on prevalence date. | Medical records from neurology departments. Contacted neurologists and psychiatrists to find additional cases. | Oct 1st 1993<br><br>Point prevalence. | 0.65 / 100,000 (0.32, 1.25) |
| Ohlmeier, 2019, Germany | At least two outpatient or inpatient ICD-10 codes for HD | 308<br><br>Incident and prevalent HD cases in insurance database. It appears that | 3,325,638<br><br>All ages.<br><br>Total population size on the database. | Institute for Applied Health Research Berlin (InGef) Research Database | 1st Jan 2015 – 31st Dec 2016<br><br>Period prevalence in German | 9.26 / 100,000 (8.28, 10.36) |

| | | | | | | |
|---|---|---|---|---|---|---|
| | | patients dying during the study period would contribute to the prevalence estimate. | | | population between 2015-2016. | |
| Panas, 2011, Greece | Symptomatic and genetic confirmation. Also included cases in relatives with a clinical diagnosis. | 594<br><br>Incident HD cases in Greece over the study period. No exclusion of deceased patients. | 10,964,020<br><br>All ages.<br><br>Total population size of Greece (unclear on what date). | Neurogenetics Laboratory | 1995 – 2008<br><br>Cumulative incidence of Greek HD patients over a 14-year period. | 5.42 / 100,000 (5, 5.87) |
| Paradisi, 2008, Venezuela | Symptomatic and genetic confirmation. | 88<br><br>(139 with expanded HTT, but only 88 symptomatic)<br><br>Incident HD cases in Venezuela over the study period. No exclusion of deceased patients. | Produced by back-calculation:<br>(n = (139 / 0.5)*100,000)<br><br>27,800,000<br><br>All ages.<br><br>Total population size of Venezuela (unclear on what date). | Laboratory of human genetics at Instituto Venezolano de Investigaciones Científicas. | 1988 – 2006<br><br>Cumulative incidence of Venezuelan HD patients over a 19-year period. | 0.32 / 100,000 (0.26, 0.39) |
| Peterlin, 2009, Slovenia | Symptomatic + CAG repeat length > 36 | 104 | 2,011,614<br><br>All ages. | Slovene Registry of patients, Genetics and Neurology | Dec 31st 2006 | 5.17 / 100,000 (4.26, 6.27) |

| | | | | | | |
|---|---|---|---|---|---|---|
| | | Total number of patients alive on prevalence date. | Total population size of Slovenia on prevalence date. | departments, Registry of Slovene Associations of HD patients. | Point prevalence. | |
| Roos, 2017, Sweden (Jamtland and Uppsala) | ICD-10 codes G10 or F02.2. | 28 (Jämtland)<br><br>17 (Uppsala)<br><br>Incident cases between 2004-2015, with deceased patients on prevalence date excluded. | 126,765 (Jämtland)<br><br>348,942 (Uppsala)<br><br>All ages.<br><br>Total population size of the regions in 2015. | Electronic medical records. | Spring 2015<br><br>Cumulative incidence of Swedish patients between 2004-2015 but with deceased patients on prevalence date excluded. | 22.1 / 100,000 (15.15, 32.06)<br><br>(Jämtland)<br><br>4.9 / 100,000 (2.98, 7.87)<br><br>(Uppsala) |
| Sackley, 2011, UK | Read code identifying the condition. | 177<br><br>Total number of patients alive on prevalence date. | 2,964,386<br><br>All ages.<br><br>Total population on database on prevalence date. | The Health Improvement Network (THIN) research database. | 2008<br><br>Point prevalence. | 5.97 / 100,000 (5.15, 6.92) |
| Shaw, 2022, Canada (Alberta) | At least two outpatient or inpatient ICD-10 codes for HD | 297<br><br>Total number of patients alive on prevalence date. | 3,183,874<br><br>>21s only.<br><br>Mid-year population size estimate for Alberta. | Province-wide administrative health data. | 2018-2019<br><br>Point prevalence. | 9.33 / 100,000 (8.32, 10.45) |
| Sipilä, 2015, Finland | ICD-9 code 3334A or ICD-10 code G10. | 114 | Produced by back-calculation: | Finnish hospital discharge register. | 31$^{st}$ Dec 2010 | 2.12 / 100,000 (1.76, 2.55) |

| | | | | | | |
|---|---|---|---|---|---|---|
| | Cases reviewed and clinical diagnosis confirmed if motor phenotype + genetic confirmation/family history. | Total number of patients diagnosed between 1987-2010 who were alive on prevalence date. | N = (114/2.12) x 100,000<br><br>5,377,358<br><br>All ages.<br><br>Appears to be total population size of Finland on or close to prevalence date. | | Cumulative incidence of Finnish patients between 1987-2010 but with deceased patients on prevalence date excluded. | |
| Sipilä, 2023, Finland | G10 diagnostic code on database. | 142<br><br>Total number of patients diagnosed between 1987-2020 who were alive on prevalence date. | 5,500,000<br><br>All ages.<br><br>Total population size on prevalence date. | Care Register of Healthcare Database | 31st Dec 2020<br><br>Cumulative incidence of Finnish patients between 1987-2020 but with deceased patients on prevalence date excluded. | 2.58 / 100,000 (2.19, 3.04) |
| Solís-Ańez, 2023, Chile | 'Motor-manifest' HD and a positive family history or >36 CAG repeats | 138<br><br>Unclear if cases included are all incident or a | 19,107,216<br><br>All ages. | Snowball sampling from patients who had visited the Centre for Movement | 2019<br><br>Point prevalence (unclear if | 0.72 / 100,000 (0.61, 0.85) |

| | | | | | | |
|---|---|---|---|---|---|---|
| | | mixture of incident/prevalent. Deaths are excluded. | Total population of Chile in 2019. | Disorders (CETRAM) | prevalent cases before 2013 included). | |
| Squitieri, 2020, Oman | Frankly symptomatic (confidence level of 4 on UDHRS) or diagnosis confirmed through genetic testing. | 41<br><br>Total number of patients alive on prevalence date. | 556,731<br><br>All ages.<br><br>Total population of Muscat region on prevalence date, | Records from the National Genetic Centre. Secondary cases actively sought through family interviews. | 2019<br><br>Point prevalence. | 7.36 / 100,000 (5.4, 10.02) |
| Squitieri, 2015, Italy (Molise) | Clinical diagnosis. | 34<br><br>Total number of patients alive on prevalence date. | 313,341<br><br>All ages.<br><br>Total population size of Molise on prevalence date. | Records from the Italian League for Research on Huntington and related diseases Foundation. Active case-finding for secondary cases were undertaken through interviews of clinicians and families, and hospital/clinic record review. | Dec 2013<br><br>Point prevalence. | 10.85 /100,000 (7.72, 15.21) |
| Sveinsson, 2012, Iceland | Symptoms + Genetic Diagnosis or positive family history | 3<br><br>Total number of patients alive on prevalence date. | 311,114<br><br>All ages.<br><br>Total population size of Iceland on prevalence date. | Healthcare records. Physicians and family members interviewed. | July 1st 2007<br><br>Point prevalence. | 0.96 / 100,000 (0.18, 2.98) |

| | | | | | | |
|---|---|---|---|---|---|---|
| Tassicker, 2008, Australia (Victoria) | Presence on disease register. | 382<br><br>Total number of patients on prevalence date – uncertainty over 58 (10 + 48) individuals who may have died prior to study date. | 4,736,000<br><br>All ages.<br><br>Total population size of Victoria on prevalence date. | Huntington disease register of Victoria | 1999<br><br>Point prevalence. | 8.07 / 100,000 (7.3, 8.92) |
| Vicente, 2021, Spain (Navarre) | Symptoms + >35 CAG repeats or positive family history. | 32<br><br>Total number of patients alive on prevalence date. | 647,554<br><br>All ages.<br><br>Total population of Navarre on prevalence date. | Administrative medical records, disability records, and the Medical Genetics Centre records. | 31st Dec 2017<br><br>Point prevalence. | 4.94 / 100,000 (3.48, 7) |
| Vishnevetsky, 2022, Peru | Genetically confirmed HD | 475<br><br>Total number of patients visiting the clinic between 2000-2018 (likely includes a mixture of incident and prevalent cases, without correcting for deaths). | 32,203,944<br><br>All ages.<br><br>Denominator data taken from world bank population estimate. For 2018. | Clinical database in Lima. | 2000-2018<br><br>Cumulative incidence of Peruvian patients between 2000-2018. 5 identified deceased JoHD patients are excluded; adult patients not | 1.46 / 100,000 (1.35, 1.61) |

| | | | | | |
|---|---|---|---|---|---|
| | | | | | corrected for those deceased. |

**Supplementary Table 2: Risk of Bias**

| Study, year, location | Appropriate sample frame or whole population? | Appropriate sampling of study participants (or whole population) | Adequate sample size* (or whole population) | Clearly defined study subjects and setting | Data analysis provided sufficient coverage of sample | Valid approach to identifying HD | Standard diagnostic approach | Appropriate statistical analysis | Adequate response rate |
|---|---|---|---|---|---|---|---|---|---|
| Alonso, 2009, Mexico (Mexico City) | 1 | 1 | 1 | 0 | NA | 1 | 1 | 0 | NA |
| Baine, 2016, South Africa | 0 | 1 | 1 | 1 | NA | 1 | 1 | 1 | NA |
| Baryshnikova, 2002, Russia (Vladimir Oblast) | 1 | 1 | 1 | 1 | NA | 1 | 1 | 1 | NA |
| Bouhouche, 2015, Morocco, Rabat-Salé | 1 | 1 | 1 | 0 | NA | 1 | 1 | 0 | NA |
| Carrasi, 2017, Italy (Ferrara) | 1 | 1 | 1 | 1 | NA | 1 | 1 | 1 | NA |
| Castilhos, 2019, Brazil (Rio Grande do Sul) | 1 | 1 | 1 | 1 | NA | 1 | 1 | 0 | NA |
| Chen, 2010, Taiwan | 1 | 1 | 1 | 1 | NA | 0 | 1 | 1 | NA |
| Cubo, 2017, Cameroon | 1 | 1 | 1 | 1 | NA | 0 | 1 | 1 | NA |

| | | | | | | | | | |
|---|---|---|---|---|---|---|---|---|---|
| Demetriou, 2017, Cyprus | 1 | 1 | 1 | 1 | NA | 1 | 1 | 0 | NA |
| Douglas + Evans, 2013, UK | 1 | 0 | 1 | 1 | NA | 0 | 1 | 1 | NA |
| Fisher, 2013, Canada (British Columbia) | 1 | 1 | 1 | 1 | NA | 1 | 1 | 1 | NA |
| Furby, 2022, UK | 0 | 0 | 1 | 1 | NA | 0 | 1 | 1 | NA |
| Furby, 2023, Sweden | 1 | 1 | 1 | 1 | NA | 0 | 1 | 1 | NA |
| Gassivaro, 1999, Malta | 1 | 1 | 1 | 1 | NA | 1 | 1 | 1 | NA |
| Gavrielov-Yusim, 2021, Israel | 0 | 0 | 0 | 1 | NA | 0 | 1 | 1 | NA |
| Gilling, 2017, Denmark | 1 | 1 | 1 | 1 | NA | 1 | 1 | 1 | NA |
| Gordon, 2016, US (Navajo Nation) | 1 | 1 | 1 | 1 | NA | 0 | 1 | 1 | NA |
| Hecimovic, 2002, Croatia | 0 | 0 | 1 | 1 | NA | 1 | 1 | 0 | NA |
| James, 1994, Wales | 1 | 1 | 1 | 1 | NA | 1 | 1 | 1 | NA |
| Kim, 2015, Korea | 1 | 1 | 1 | 1 | NA | 0 | 1 | 1 | NA |
| Kounidas, 2021, Scotland | 1 | 1 | 1 | 1 | NA | 1 | 1 | 0 | NA |

| | | | | | | | | | |
|---|---|---|---|---|---|---|---|---|---|
| Lee, 2023, South Korea | 1 | 1 | 1 | 1 | NA | 0 | 1 | 1 | NA |
| McCusker, 2000, Australia (New South Wales) | 1 | 1 | 1 | 1 | NA | 1 | 1 | 1 | NA |
| Morrison, 2010, Northern Ireland | 1 | 1 | 1 | 1 | NA | 1 | 1 | 0 | NA |
| Muroni, 2020, Italy (Cagliari and South Sardinia) | 1 | 1 | 1 | 1 | NA | 1 | 1 | 1 | NA |
| Nakashima, 1993, Japan (San-In Area) | 1 | 1 | 1 | 1 | NA | 1 | 1 | 1 | NA |
| Ohlmeier, 2019, Germany | 1 | 0 | 1 | 1 | NA | 0 | 1 | 1 | NA |
| Panas, 2011, Greece | 1 | 1 | 1 | 1 | NA | 1 | 1 | 1 | NA |
| Paradisi, 2008, Venezuela | 1 | 1 | 1 | 1 | NA | 1 | 1 | 0 | NA |
| Peterlin, 2009, Slovenia | 1 | 1 | 1 | 1 | NA | 1 | 1 | 1 | NA |
| Roos, 2017, Sweden (Jamtland and Uppsala) | 1 | 1 | 1 | 1 | NA | 1 | 1 | 1 | NA |
| Sackley, 2011, UK | 1 | 1 | 1 | 1 | NA | 1 | 1 | 1 | NA |

| | | | | | | | | | |
|---|---|---|---|---|---|---|---|---|---|
| Shaw, 2022, Canada (Alberta) | 1 | 0 | 1 | 1 | NA | 0 | 1 | 0 | NA |
| Sipilä, 2015, Finland | 1 | 1 | 1 | 0 | NA | 1 | 1 | 1 | NA |
| Sipilä, 2023, Finland | 1 | 1 | 1 | 1 | NA | 0 | 0 | 1 | NA |
| Solís-Ańez, 2023, Chile | 0 | 0 | 1 | 1 | NA | 1 | 1 | 1 | NA |
| Squitieri, 2020, Oman | 1 | 1 | 1 | 1 | NA | 1 | 1 | 1 | NA |
| Squitieri, 2015, Italy (Molise) | 1 | 1 | 1 | 1 | NA | 1 | 1 | 1 | NA |
| Sveinsson, 2012, Iceland | 1 | 1 | 1 | 1 | NA | 1 | 1 | 1 | NA |
| Tassicker, 2008, Australia (Victoria) | 1 | 1 | 1 | 1 | NA | 1 | 1 | 1 | NA |
| Vicente, 2021, Spain (Navarre) | 1 | 1 | 1 | 1 | NA | 1 | 1 | 1 | NA |
| Vishnevetsky, 2022, Peru | 1 | 1 | 1 | 1 | NA | 0 | 1 | 0 | NA |

*Adequate sample size calculated as: >2,268,042

**Supplementary Table 3: Meta-regression Variables**

| Study, year, location | Case Ascertainment | Region | Median Age (source) | Healthcare Access and Quality Index (HAQI) Score | Years after 1993 |
|---|---|---|---|---|---|
| Alonso, 2009, Mexico (Mexico City) | Passive | South America | 24.3 (Database earth) | 60.5 | 15 |
| Baine, 2016, South Africa | Administrative | Africa | 25 (Database earth) | 52 | 21 |
| Baryshnikova, 2002, Russia (Vladimir Oblast) | Administrative | Europe | 35.4 (Database earth) | 59.7 | 6 |
| Bouhouche, 2015, Morocco, Rabat-Salé | Administrative | Africa | 26.1 (Database earth – median age for Morocco as a whole applied) | 61.3 | 21 |
| Carrasi, 2017, Italy (Ferrara) | Administrative | Europe | 44 (Database earth – median age for Italy as a whole applied) | 88.7 | 21 |
| Castilhos, 2019, Brazil (Rio Grande do Sul) | Administrative | South America | 30.7 (Database earth) | 64.9 | 23 |
| Chen, 2010, Taiwan | Administrative | Asia | 33.8 (Worldometers – median age for 2005 applied) | 73.6 | 14 |
| Cubo, 2017, Cameroon | Administrative | Africa | 17 (Database earth) | 44.4 | 21 |
| Douglas + Evans, 2013, UK | Administrative | Europe | 38.2 (Database earth) | 82.7 | 17 |

| | | | | | |
|---|---|---|---|---|---|
| Demetriou, 2017, Cyprus | Administrative + Active Case Finding | Europe | 34.9 (Database earth) | 85.3 | 22 |
| Fisher, 2013, Canada (British Columbia) | Administrative + Active Case Finding | North America | 39.1 (Database earth) | 86.3 | 19 |
| Furby, 2022, UK | Administrative | Europe | 39.1 (Database earth) | 84.6 | 25 |
| Furby, 2023, Sweden | Administrative | Europe | 39.6 (Database earth) | 90.5 | 25 |
| Gassivaro, 1999, Malta | Administrative | Europe | 33.3 (Database earth) | 76.8 | 0 |
| Gavrielov-Yusim, 2021, Israel | Administrative | Asia | 28.9 (Database earth) | 85.5 | 25 |
| Gilling, 2017, Denmark | Administrative | Europe | 40.4 (Database earth) | 85.7 | 22 |
| Gordon, 2016, US (Navajo) | Administrative | North America | 29.9 (Community Survey of Navajo Nation) | 81.3 | 18 |
| Hecimovic, 2002, Croatia | Administrative | Europe | 38.43 (Database earth) | 73.4 | 9 |
| James, 1994, Wales | Administrative | Europe | 35.4 (Database earth – median age for UK as a whole applied) | 76.6 | 1 |
| Kim, 2015, Korea | Administrative | Asia | 38.56 (Database earth) | 85.8 | 20 |
| Kounidas, 2021, Scotland | Administrative | Europe | 39.2 (Database earth – median age for UK as a whole applied) | 84.6 | 27 |
| Lee, 2023, South Korea | Administrative | Asia | 42.2 (Database earth) | 85.8 | 26 |

| | | | | | |
|---|---|---|---|---|---|
| McCusker, 2000, Australia (New South Wales) | Administrative + Active Case Finding | Europe | 33 (Database earth – median age for Australia as a whole applied) | 80.8 | 3 |
| Morrison, 2010, Northern Ireland | Administrative | Europe | 36.8 (Database earth – median age for UK as a whole applied) | 78.4 | 8 |
| Muroni, 2020, Italy (Cagliari and South Sardinia) | Administrative + Active Case Finding | Europe | 45.2 (Database earth – median age for Italy as a whole applied) | 88.7 | 24 |
| Nakashima, 1993, Japan (San-In Area) | Passive | Asia | 38.2 (Database earth) | 80.7 | 0 |
| Ohlmeier, 2019, Germany | Administrative | Europe | 44.9 (Database earth) | 86.4 | 23 |
| Panas, 2011, Greece | Administrative | Europe | 39.8 (Database earth) | 85.2 | 15 |
| Paradisi, 2008, Venezuela | Administrative | South America | 23.7 (Database earth) | 62.3 | 13 |
| Peterlin, 2009, Slovenia | Administrative | Europe | 39.1 (Database earth) | 79.8 | 13 |
| Roos, 2017, Sweden (Jamtland and Uppsala) | Administrative | Europe | 39.9 (Database earth – median age for Sweden as a whole applied) | 90.5 | 22 |
| Sackley, 2011, UK | Administrative | Europe | 37.9 (Database earth) | 82.7 | 15 |
| Shaw, 2022, Canada (Alberta) | Administrative | North America | 39.8 (Database earth – median age for Canada as a whole applied) | 87.6 | 26 |
| Sipilä, 2015, Finland | Administrative | Europe | 41 (Database earth) | 87.2 | 17 |
| Sipilä, 2023, Finland | Administrative | Europe | 39.5 (Database earth) | 90.5 | 27 |

| | | | | | |
|---|---|---|---|---|---|
| Solís-Ańez, 2023, Chile | Administrative | South America | 34.3 (Database earth) | 76 | 26 |
| Squitieri, 2020, Oman | Administrative + Active Case Finding | Asia | 28.8 (Database earth) | 77.1 | 20 |
| Squitieri, 2015, Italy (Molise) | Administrative + Active Case Finding | Europe | 43.6 (Database earth – median age for Italy as a whole applied) | 88.7 | 26 |
| Sveinsson, 2012, Iceland | Administrative + Active Case Finding | Europe | 33.6 (Database earth) | 89.8 | 14 |
| Tassicker, 2008, Australia (Victoria) | Administrative | Oceania | 35.1 (Australian Demographics for Victoria) | 83.7 | 6 |
| Vicente, 2021, Spain (Navarre) | Administrative + Active Case Finding | Europe | 42.4 (Database earth – median age for Spain as a whole applied) | 89.6 | 24 |
| Vishnevetsky, 2022, Peru | Administrative | South America | 27.43 (Database earth) | 69.6 | 25 |

Healthcare Access and Quality Index (HAQI) scores are all taken from the same website, with the closest year available to prevalence date applied. Please note the website is now only available as an archived version.

### Sensitivity Analysis: Supplementary Forest Plots

<u>World Bank Regions</u>

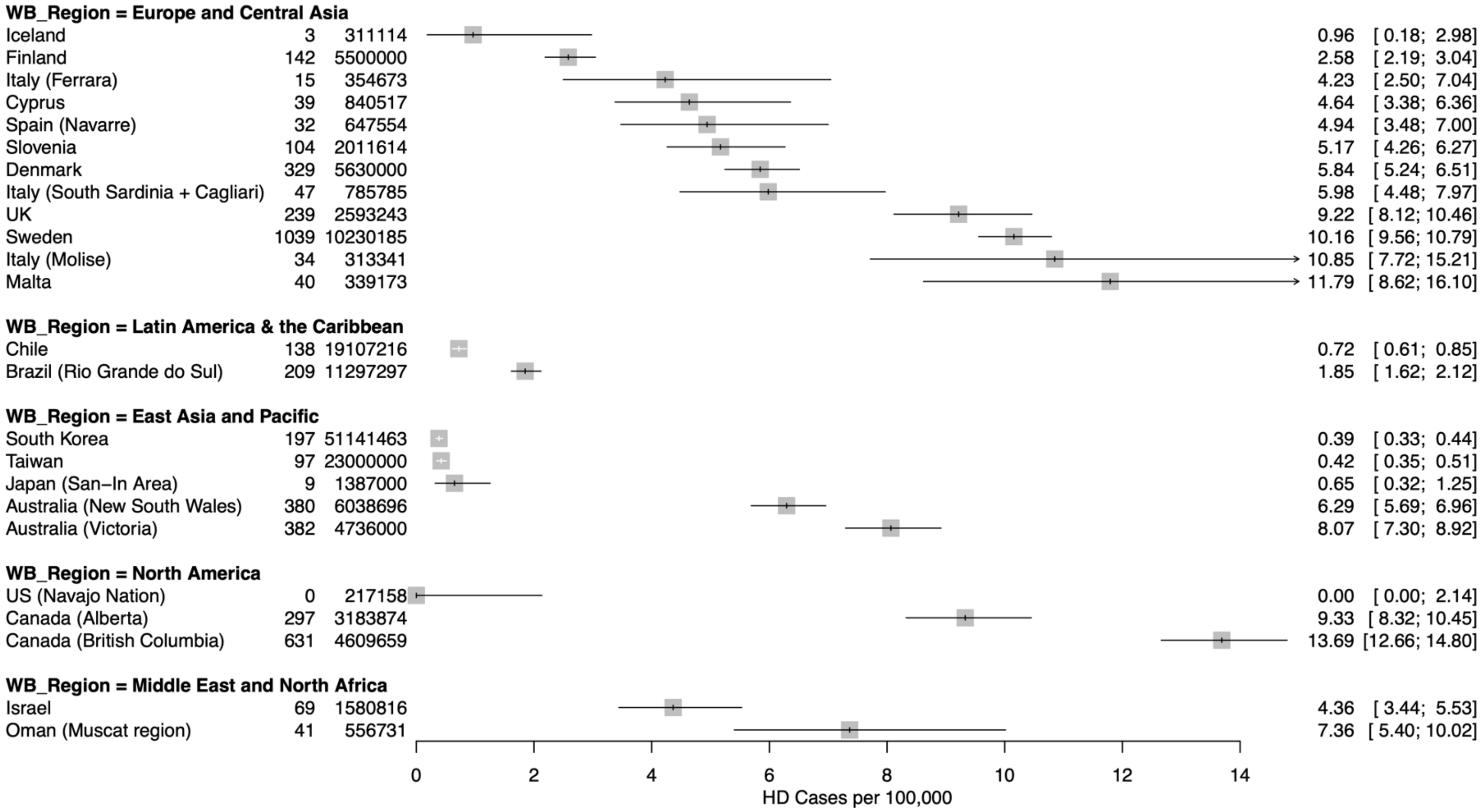

Excluding studies based on administrative codes alone

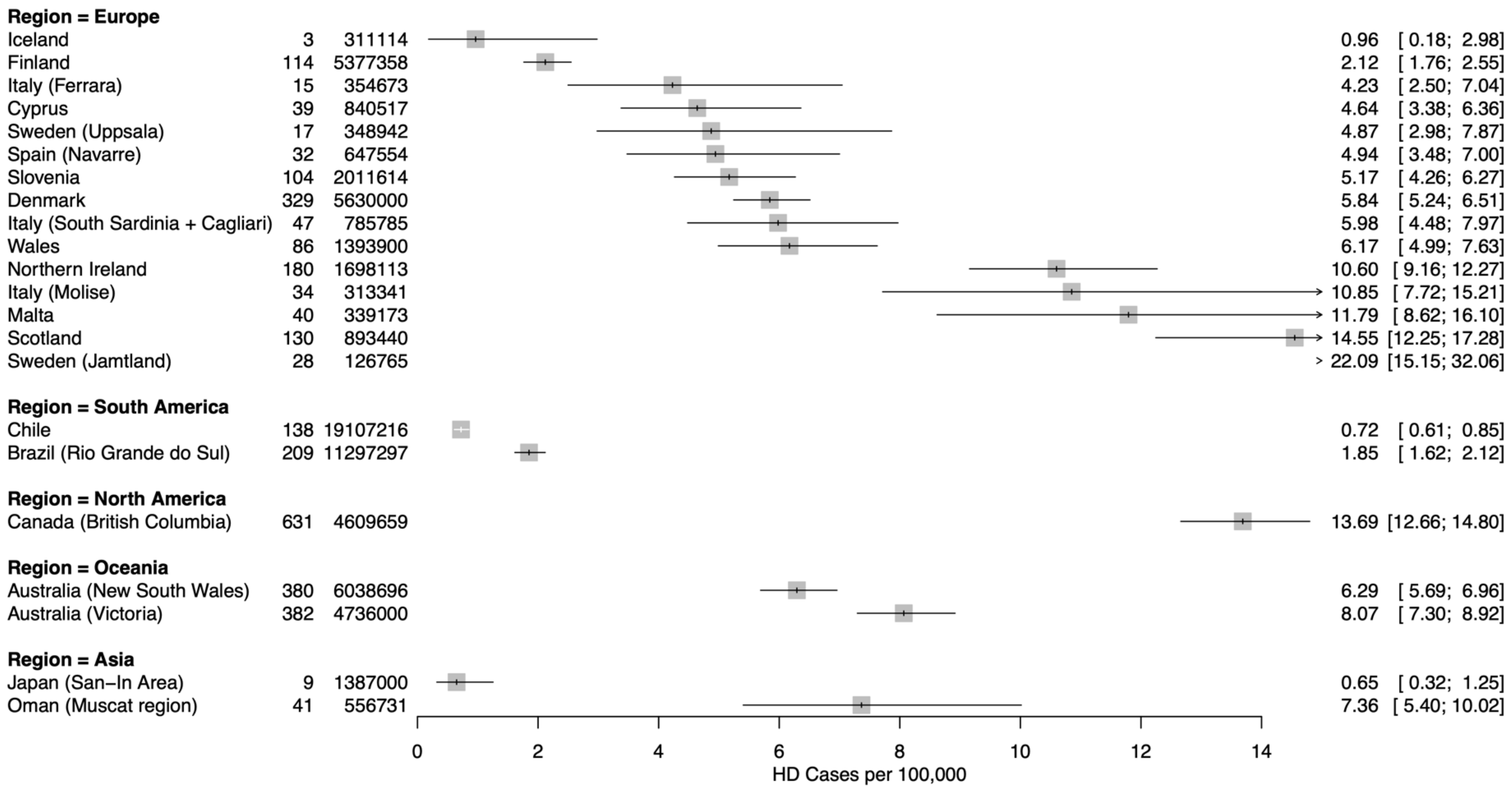

**Sensitivity Analysis: Supplementary Meta-Regression Tables**

| **World Bank Region** | | | |
|---|---|---|---|
| Variable | Exponentiated Coefficients (CI, 95%) | Standard Error | P - value |
| Region | | | |
| Europe and Central Asia | 1 (ref) | NA | NA |
| East Asia and Pacific | 0.26 (0.05, 1.21) | 0.73 | 0.081 |
| Latin America & the Caribbean | 0.44 (0.03, 6.85) | 1.29 | 0.536 |
| North America | 1.19 (0.31, 4.56) | 0.63 | 0.792 |
| Middle East and North Africa | 3.3 (0.31, 35.57) | 1.12 | 0.301 |
| Median age | 1.09 (0.93, 1.27) | 0.07 | 0.273 |
| Year post-1993 | 0.96 (0.88, 1.04) | 0.04 | 0.267 |
| Case Ascertainment Method | | | |
| Passive | 1 (ref) | NA | NA |
| Passive + Active Case Finding | 1.16 (0.4, 3.39) | 0.5 | 0.766 |
| Healthcare Access and Quality Index (HAQI) score | 1 (0.88, 1.13) | 0.06 | 0.993 |
| Constant | <0.001 | 4.65 | 0.019 |
| $R^2$ | 0.43 | | |

| **Excluding Administrative Code Based Studies** | | | |
|---|---|---|---|
| Variable | Exponentiated Coefficients (CI, 95%) | Standard Error | P - value |
| Region | | | |
| Europe | 1 (ref) | NA | NA |
| Asia | 0.21 (0.07, 0.61) | 0.48 | 0.007 |
| South America | 0.02 (<0.01, 0.08) | 0.66 | <0.001 |
| Oceania | 1.68 (0.52, 5.41) | 0.54 | 0.353 |
| Median age | 1 (0.91, 1.11) | 0.05 | 0.949 |
| Year post-1993 | 1.07 (1.02, 1.11) | 0.02 | 0.004 |
| Case Ascertainment Method | | | |
| Passive | 1 (ref) | NA | NA |
| Passive + Active Case Finding | 0.88 (0.46, 1.67) | 0.3 | 0.665 |
| Healthcare Access and Quality Index (HAQI) score | 0.88 (0.81, 0.96) | 0.04 | 0.008 |
| Constant | 0.83 | 2.8 | 0.947 |
| $R^2$ | 0.69 | | |
